# Expansion of the HSV-2-specific T cell repertoire in skin after immunotherapeutic HSV-2 vaccine

**DOI:** 10.1101/2022.02.04.22270210

**Authors:** Emily S. Ford, Alvason Li, Kerry J. Laing, Lichun Dong, Kurt Diem, Lichen Jing, Krithi Basu, Mariliis Ott, Jim Tartaglia, Sanjay Gurunathan, Jack L. Reid, Matyas Ecsedi, Aude G. Chapuis, Meei-Li Huang, Amalia S. Magaret, Christine Johnston, Jia Zhu, David M. Koelle, Lawrence Corey

## Abstract

The skin at the site of HSV-2 reactivation is enriched for HSV-2-specific T cells. To evaluate whether an immunotherapeutic vaccine could elicit skin-based memory T cells, we studied skin biopsies and HSV-2-reactive CD4^+^ T cells from peripheral blood mononuclear cells (PBMCs) by T cell receptor β (*TRB*) sequencing before and after vaccination with a replication-incompetent whole virus HSV-2 vaccine candidate (HSV529). The representation of HSV-2-reactive CD4^+^ *TRB* sequences from PBMCs in the skin *TRB* repertoire increased after the first vaccine dose. We found sustained expansion after vaccination of unique, skin-based T-cell clonotypes that were not detected in HSV-2-reactive CD4^+^ T cells isolated from PBMCs. In one participant a switch in immunodominance occurred with the emergence of a T cell receptor (TCR) αβ pair after vaccination that was not detected in blood. This TCRαβ was shown to be HSV-2-reactive by expression of a synthetic TCR in a Jurkat-based NR4A1 reporter system. The skin in areas of HSV-2 reactivation possesses an oligoclonal *TRB* repertoire that is distinct from the circulation. Defining the influence of therapeutic vaccination on the HSV-2-specific *TRB* repertoire requires tissue-based evaluation.

## INTRODUCTION

While antivirals for HSV-2 have been in widespread use for over 35 years, HSV-2 infection continues to be highly prevalent globally (*1–3*). As such, development of novel methods to curtail HSV-2 reactivation and transmission are needed. T cell immune responses are associated with severity and reactivation frequency of mucosal HSV infections (*4*, *5*), contributing to interest in the development of immunotherapeutic approaches to control HSV-2 reactivation. Although recombinant HSV-2 protein-based vaccines have been tested, and one demonstrated a partial but significant reduction in the rate of subclinical HSV-2 reactivation among seropositive adults (*5–8*), correlation between humoral or cellular markers of HSV-2 immunogenicity and vaccine response has not been demonstrated (*5*, *8–11*). Likewise, T cell responses in peripheral blood mononuclear cells (PBMCs) were not associated with efficacy in studies of peptide-based vaccines (*12*, *13*). In a Phase 1 study of HSV529, a replication-deficient HSV-2 whole virus vaccine, 46% of HSV-2-seropositive participants had a detectable increase in IFNγ expression in CD4^+^ T cells from PBMCs (*14*). There was minimal (<10%) change in IFNγ expression from CD8^+^ T cells over the course of the vaccine trial.

Except during acute infection, HSV-specific CD4^+^ T cells comprise only a small percentage of circulating CD4^+^ T cells (*15*). T cell production of IFNγ or other cytokines in response to whole virus or peptide-based libraries is typically used to determine HSV specificity or activity (*16–19*). In a study of 40 HSV-2-seropositive individuals during clinical quiescence, 0.04-3.77% (median 0.31%) of CD4^+^ T cells from PBMCs expressed IFNγ after HSV-2 stimulation in vitro (*20*). As a surrogate marker of activation, HSV-specific CD4^+^ and CD8^+^ T cells can express CD137 after in vitro exposure to antigen and antigen-presenting cells in activation-induced marker (AIM) assays (*21*). Cytokine production after cognate antigen exposure remains the gold standard to identify T cells with HSV specificity used in studies of vaccine response.

Data from animal models, corroborated by mathematical modeling of human data and observational human studies, have supported the importance of skin-resident memory T cells (T_RM_) in the adaptive immune responses to HSV-2 reactivation (*22–27*). These cells are distinct from skin-migratory T cells by phenotype, skin ingress and egress markers, and spatial localization (*27–29*). A potential explanation for the lack of association between clinical benefit and measured immune response in immunotherapeutic vaccine trials is that the immune correlates of efficacy may lie in alteration of the immune responses in the genital or mucosal skin, rather than the response detected in circulating PBMCs.

To investigate whether a 3-dose vaccination series with HSV529 could elicit changes in the skin-based T cell response, we performed T cell receptor (TCR) β chain (*TRB*) repertoire sequencing and quantitation of the number of copies of each unique *TRB* sequence (clonotype) of genital skin biopsies taken during clinical quiescence from the site of previous symptomatic HSV-2 reactivation before and after each dose. The *TRB* repertoire of HSV-2-reactive CD4^+^ T cells from PBMCs was also obtained to allow for extrapolation of probable HSV-2-specificity among skin-identified clonotypes. CD4^+^ T cells were selected for comparison due to their observed expansion in the preceding phase 1 clinical trial (clinicaltrials.gov, NCT02571166). We found that an immunotherapeutic vaccination elicits skin-based immune responses including expansion of HSV-2-specific T-cell clonotypes. Expansion in HSV-2-specific CD4^+^ clonotypes targeting many varied HSV-2 epitopes was observed in blood. We also found sustained expansion after vaccination of unique, skin-based T cell clonotypes that were not detected in HSV-2-reactive CD4^+^ T cells isolated from PBMCs.

## RESULTS

### Vaccination does not alter T cell abundance or localization at sites of HSV-2 reactivation

To determine whether an immunotherapeutic vaccine against HSV-2 might alter the T cell repertoire at the site of HSV-2 reactivation, nine persons between the ages of 32 and 54 years with a known history of symptomatic HSV-2 (median duration 11.9 years) were enrolled (**Fig. 1a, Tables S1, S2**) (clinicaltrials.gov, NCT02571166). Vaccine doses were given at days 0, 30, and 180 and biopsies were performed at days 0, 10, 30, 40, 180, and 190 (**Fig. 1a**). Seven participants completed the entire three-dose, seven-biopsy study protocol; two completed biopsies through day 40.

**Fig. 1.**
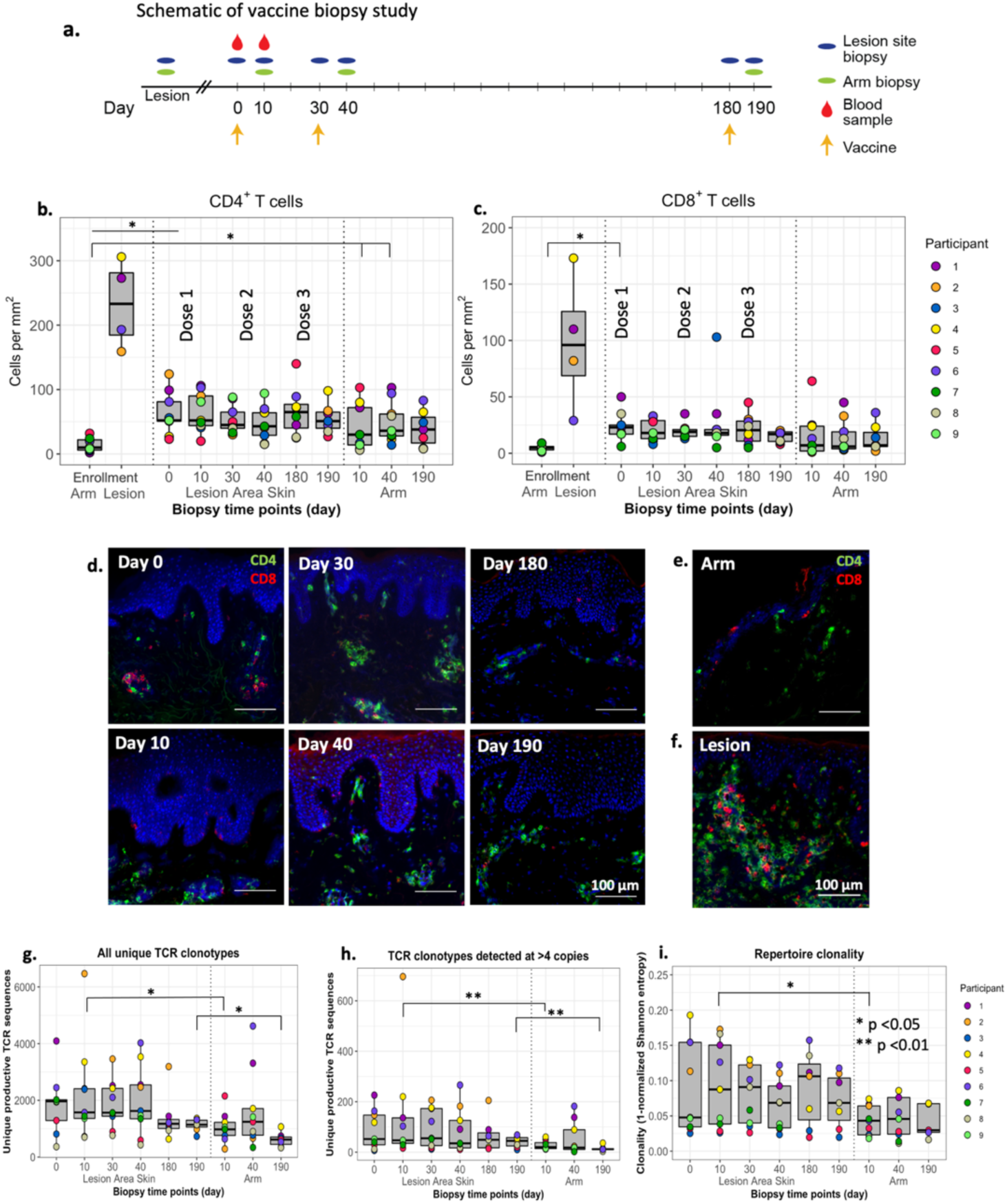
Number, fold change, and clonality of TCR clonotypes in HSV lesion site and arm biopsies before and after vaccination. (**a**) Schematic of vaccine study timeline and procedures. CD4^+^ (**b**) and CD8^+^ (**c**) T-cell densities from biopsies from control skin at the time of enrollment and from the site of a symptomatic lesion (N=4) are shown in comparison to HSV lesion site and control skin biopsies over the course of a 3-dose vaccine trial in nine vaccine recipients. Each dot represents the mean of three counted sections in a single participant. Median and interquartile range are shown in grey. (**d**) Representative 10x micrographs of CD4^+^ (green) and CD8^+^ (red) T-cell density by IFA in HSV lesion site biopsies at specified time points before and after vaccination. CD4^+^ and CD8^+^ T-cell IFA from (**e**) control skin and (**f**) lesion site during a symptomatic HSV-2 outbreak. All images are from P4. White bars are 100μm. (**g**) Total and (**h**) number of TCRβ clonotypes detected at >4 copies are shown from the HSV lesion site and arm biopsies. (**h**) Clonality calculated from Shannon entropy of the TCR repertoire from each sample. Each dot represents a single participant. Median and interquartile range are shown in grey. For comparisons, *p <0.05, **p <0.01 by paired Wilcoxon.

To determine whether vaccination influenced the size of the skin-based T cell repertoire, the density of CD4^+^ and CD8^+^ T cells resident during an active lesion, clinical quiescence and before and after each vaccination dose were enumerated in biopsies from the lesion-area skin. T cell immunofluorescence (IF) showed that vaccination was not associated with an increase in CD4^+^ or CD8^+^ T cell density at the lesion site (**Fig. 1b, c**). We also did not observe differences in CD4^+^ or CD8^+^ T cell spatial localization at either the lesion site or arm by visual inspection, which were scattered in the upper dermis, dermal epidermal junction (DEJ), or in clusters as is typically observed (**Fig. 1d, e, f**) (*30*, *31*). The density of total CD4^+^ and CD8^+^ T cell infiltration was lower in the arm at enrollment compared to the lesion area during clinical quiescence (day 0) (**Fig. 1b, c**). Perhaps related to the site of vaccination, the number of CD4^+^ T cells in the arm biopsy was significantly higher after dose 1 and 2 (day 10 and 40) compared to before vaccination (14.0 vs. 40.0 and 49.8 cells/mm^2^, p = 0.02 and 0.01, respectively), but the number of CD8^+^ T cells did not change significantly. Four participants had genital HSV-2 recurrences (lesions) in the three months before vaccination. All biopsies from these lesions demonstrated the massive lymphocyte infiltration typically associated with symptomatic reactivation (**Fig. 1f**).

### The *TRB* repertoire in lesion-area skin is larger and more diverse than in the arm

We performed bulk *TRB* sequencing from skin to visualize the *TRB* repertoire in lesion-area skin in comparison to the arm. *TRB* sequencing from lesion site biopsies taken during clinical quiescence, just before vaccination (day 0), identified a median of 1,966 unique clonotypes and 52 clonotypes identified at >4 copies per participant. Vaccination did not result in a significant increase in the median number of unique clonotypes in the lesion area per participant or the number of unique clonotypes detected at >4 copies (**Fig. 1g, h**). More clonotypes were detected at >4 copies in HSV lesion sites than in the arm at two of three sampled timepoints (day 10 and 190, p < 0.01) (**Fig. 1h**). *TRB* repertoire clonality, defined as the inversion of Shannon entropy (a comparative measure of the number and frequency of unique TRB sequences in each sample) (*32*), was greater in lesion site than arm biopsies at day 10 (p = 0.03), but did not change in the lesion site after any of the three vaccinations (**Fig 1i**).

### Vaccination elicited HSV-2-reactive CD4^+^ T cells in blood

To obtain *TRB* sequences to perform clonal tracking of a known population of HSV-2 reactive CD4^+^ T cells, HSV-2 reactive CD4^+^ T cells were isolated from PBMCs before and after the first vaccine dose (see Methods). At day 0, a median of 30 unique clonotypes per participant were identified (range 25-51). After vaccination, the median number of unique HSV-2-reactive clonotypes increased 8-fold to a median of 237 clonotypes/participant (range 46-1360, unpaired Wilcoxon p = 0.002 excluding participant 4 (P4), whose cells underwent ex vivo expansion prior to sequencing) (**Table S3**). Almost all of the 233 HSV-2-reactive clonotypes in PBMC at day 0 were detected at single copy, whereas at day 10, clonotypes were found at >2 copies in 6 of 9 participants, suggesting that an increase in detectable HSV-2-reactive T cells in the blood after vaccination with HSV529 comes from expansion of relatively low abundance clonotypes. The low copy number of each unique T cell sequence, particularly from day 0, is not unexpected because HSV-2-reactive T cells were not expanded in vitro prior to sequencing (**Fig. S1**).

### Almost half of vaccine-elicited HSV-2-reactive T cells from PBMCs detected in skin were present in skin prior to vaccination

We next mapped the frequency of detection and persistence of these PBMC-identified HSV-2-reactive CD4^+^ T cell clonotypes in *TRB* repertoires from lesion area and control site biopsies. Prevalent clonotypes were defined as clonotypes that had been detected prior to the initiation of the vaccine series, whereas elicited clonotypes were defined as those only detected after initiation of vaccination. Of 205 HSV-2-reactive clonotypes detected in PBMCs from all 7 tested participants at day 0, 16 (7.8%, in 4 participants) HSV-2-reactive blood clonotypes were detected in any skin biopsy from day 0 to day 190, and seven (in 3 participants) of the 16 (43%) were detected at day 0. Three participants did not have day 0 blood clonotypes detected in skin. Eight clonotypes from blood at day 0 were detected in the active lesion biopsies from 4 participants. (**Fig. 2a, b, Tables S3, S4a-c**). Ten days after dose 1, 2212 HSV-2-reactive clonotypes were detected in PBMCs; only five (0.1%) had also been detected in the day 0 blood sample. 447 of 2212 (20%) were detected in lesion-area skin from day 0 to day 190, with 203 of those 447 (45%) detected in skin at day 0 (from 8 of 9 participants) (**Figure 2c, d, Tables S3, S4a, b**), and 247 of 1920 had been detected at the time of active lesion in the 4 participants where that biopsy was available (**Table S4c**). Thus, 45% of skin-detected and 9% of all vaccine-elicited clonotypes from PBMCs were present as resident T cells in tissue at study onset (day 0) (**Fig. 2d**, P4 **Fig. 2e**). Overlap between the arm at any time point and the day 0 or day 10 HSV-2-reactive clonotypes from blood was low, ranging from 0-5 clonotypes per person from the day 0 sample and 0-25 clonotypes per person post vaccination (**Table S4**). In summary, vaccination increased the number of detectable, unique HSV-2-reactive CD4+ T cell clonotypes in blood (205 to 2212 clonotypes across all participants). Of vaccine-elicited clonotypes, approximately 20% were detected in skin biopsies at any point and nearly half of these had been present in genital area skin at the time of the initiation of the vaccine series.

**Fig. 2.**
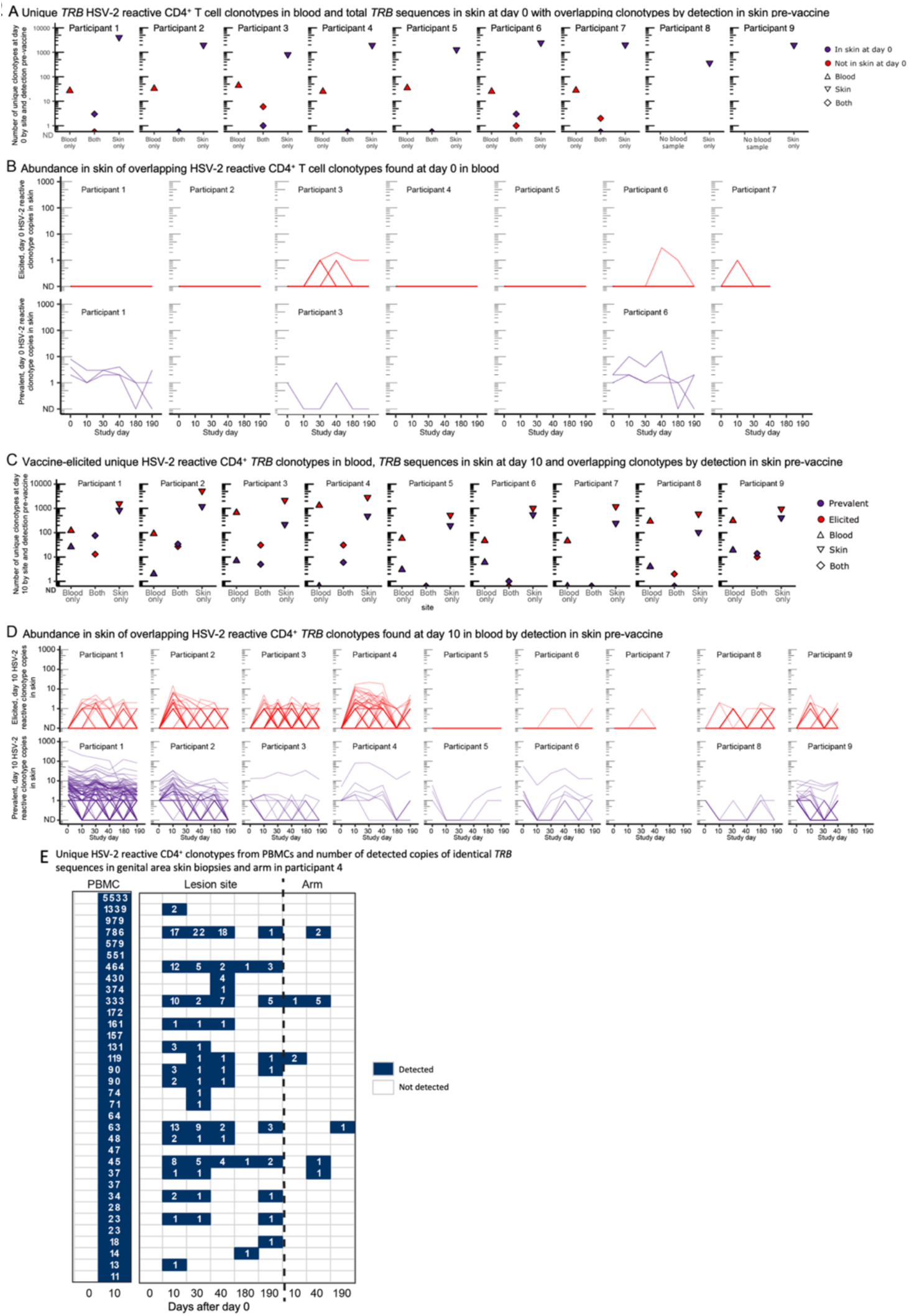
Overlap of HSV-2-reactive CD4^+^ T cells from PBMCs in skin biopsies by clonal tracking before and after initiation of vaccination series. (**a**) Unique *TRB* sequences detected only in blood at day 0 (‘blood only’), clonotypes from blood at day 0 also detected at any time in skin (‘both’) and unique clonotypes in skin at day 0 (‘skin only’) and (b) the longitudinal detection of each of the overlapping (‘both’) clonotypes in lesion-area skin over time, by whether they were observed before or after vaccination. Each line represents a unique clonotype. “ND” is not detected at that time point. (**c**) Unique *TRB* sequences detected in blood and skin at day 10 by detection in one or both sites and (**d**) the longitudinal detection of each of the overlapping clonotypes in lesion-area skin over time. Each line represents a unique clonotype. “ND” is not detected at that time point. Shading of both histograms and longitudinal graphing represents whether clonotypes were observed before (**purple**) or only after (**red**) vaccination. (**e**) All HSV-2-reactive CD4^+^ T cells from PBMCs identified in P4 at day 10 at >10 copies and their representation in serial tissue biopsies after immunotherapeutic vaccination. Empty boxes indicate no detection. Each horizontal row is a distinct nucleotide sequence. Quantitation of PBMC copy number at day 10 (but not day 0) in this participant is influenced by T cell ex vivo expansion prior to stimulation and sequencing (**Fig. S1**, method 2).

### Persistence and expansion of skin-based clonotypes

We next characterized the skin-based repertoire over time to determine how the relative abundance of clonotypes in skin changed over the course of vaccination. To narrow analysis to clonotypes of greatest interest, we identified and ranked the skin-based clonotypes with the greatest expansion (≥6-fold increase in number of copies) after the first vaccine dose (from day 0 to day 10) as “tissue-expanders” and examined their durability in serial biopsies from lesion-area skin, and detection in serial biopsies from the arm and in HSV-2-reactive CD4^+^ T cells from PBMCs. Prevalent clonotypes that increased ≥6-fold after dose 1 at the lesion site were infrequently detected in blood and did not undergo similar expansion in the arm (P4, **Fig. 3a**, all others **Fig. S2**). Clonotype trajectories of prevalent clonotypes increasing by ≥6-fold over dose 1 are shown in all participants in **Fig. 3b**. **Fig. 3c** shows elicited clonotypes that were newly identified at 6 copies or greater at day 10 after dose 1. In both **Fig. 3b** and **3c**, clonotypes identified as HSV-2 reactive CD4^+^ T cells from PBMCs are shown in red. The longitudinal detection and CDR3 amino acid sequence of each tissue-expanding clonotype over dose 1 in all participants is shown in **Fig. S2a, b** and **Table S5a,b**. The number of prevalent and elicited expanding clonotypes in each participant is summarized in **Table 1**. Among all participants, there was more expansion (by number of clonotypes increasing by ≥6 fold) in the lesion area biopsies from day 0 to day 10 (median 11, range 1-34) than in the arm biopsies from day 10 to 190 (median 0, range 0-4, p = 0.01 by paired Wilcoxon) and day 40 to 190 (median 0, range 0-3, p = 0.01). Expansion in lesion-area biopsies was greatest after dose 1 and waned after doses 2 and 3 (**Fig. 3d, Table S5c**). There were a similar number of prevalent and elicited clonotypes expanding after each dose (**Fig. 3d**), though expanding clonotypes represented a greater proportion of prevalent than elicited clonotypes (**Table 1**). Among 535 prevalent and elicited clonotypes expanding in tissue after dose 1, 21 unique clonotypes were detectable as HSV-2 reactive CD4+ T cells in blood, from only 2 participants (**Fig. 3b, c, Table 1**).

**Fig. 3.**
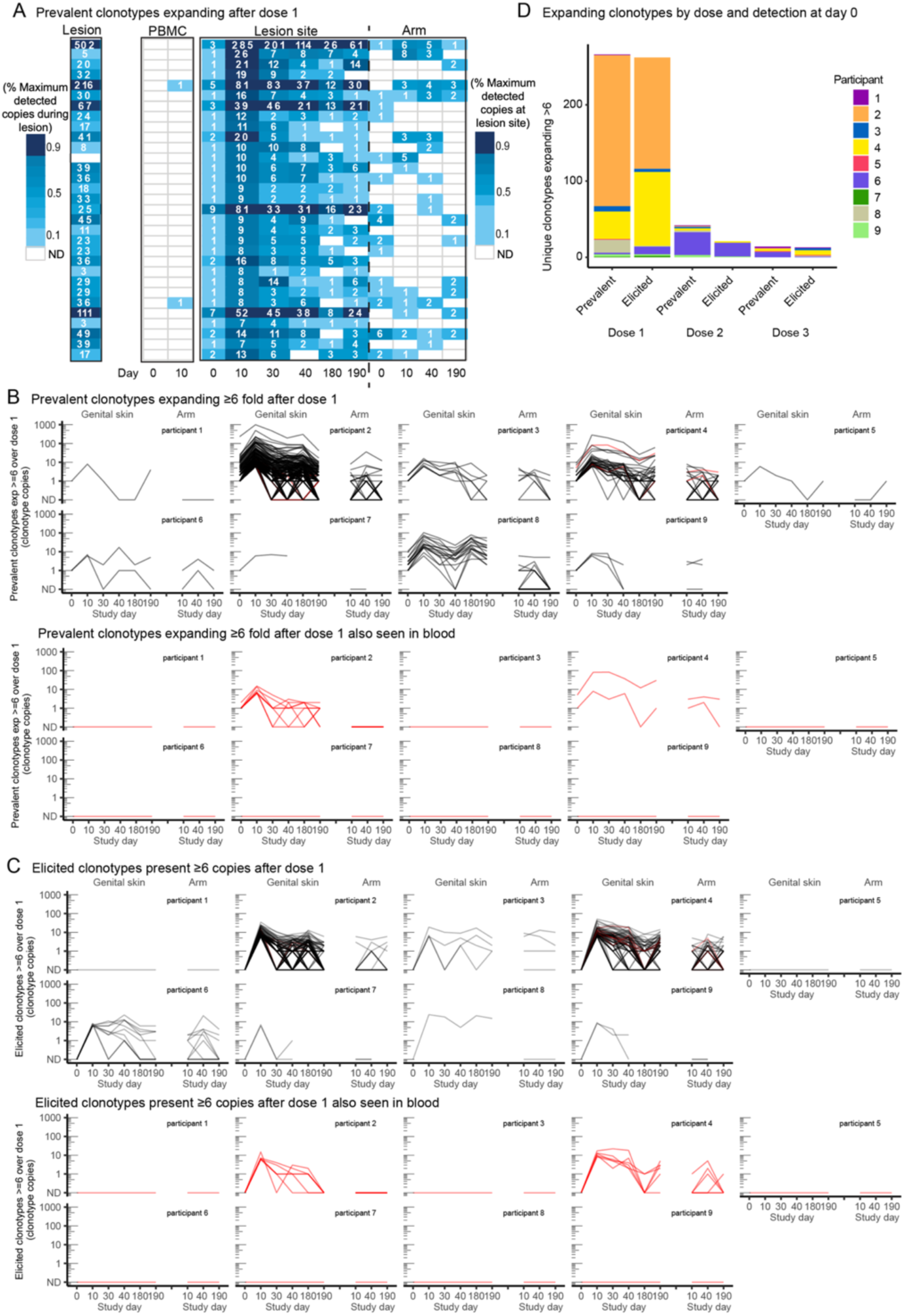
Prevalent and elicited clonotypes in HSV-2-enriched CD4+ T cells from PBMCs (“PBMC”), healed lesion site (“HSV lesion-area skin”), and arm biopsy expanding or detected at high copy number. (**a**) Prevalent clonotypes detected in P4, by fold increase over dose 1. Each row represents a single clonotype (by nucleic acid sequence). Co-detection of clonotypes in HSV-2-enriched CD4+ T cells from PBMCs, active lesion, quiescent lesion-area skin and arm skin is compared at each of the time points. Number of copies detected in PBMCs from P4 at day 10 reflect ex vivo expansion prior to sequencing and are denoted by # (see Methods). Blank boxes indicate lack of detection. (**b**) Prevalent and (**c**) elicited clonotypes either expanding 26 fold after dose 1 or present after dose 1 (day 10) at 26 fold above a single copy and their longevity in lesion area tissue over the course of the vaccine trial (left), with the corresponding abundance in the arm control (right). ND = not detected. Highly prevalent clonotypes that expanded after the first vaccine dose are seen to persist in tissue throughout the vaccine trial. Color indicates whether the nucleotide sequences were detected in blood (black indicates a tissue-only sequence, red was also seen in blood). Prevalent expanded clonotypes are of greater abundance than elicited clonotypes. Most in-tissue expansion was not from clonotypes observed in blood. (**d**) Number of clonotypes expanding by 26 fold after each dose in the lesion-area skin by detection prior to initiation of vaccine series (prevalent) vs clonotypes only seen after initiation of vaccination (elicited).

**Table 1.** Number of unique clonotypes, expansion after vaccine doses, detection in HSV-2 reactive CD4+ T cells from blood and tissue residency of prevalent and elicited clonotypes in skin.

|  | Person | Unique skin clonotypes | Detected in blood and skin (%)<sup>c</sup> | Expanders (≥6) |  |  | Total unique expanders | Dose 1 expanders detected in blood | Resident (>70% biopsies) (%)<sup>d</sup> | Resident expanders (% expanders) | Residents detected in blood (%) |
| --- | --- | --- | --- | --- | --- | --- | --- | --- | --- | --- | --- |
|  |  |  |  | 1 | 2 | 3 |  |  |  |  |  |
| Prevalent<sup>a</sup> | 1 | 4092 | 103 (2.5) | 1 | 1 | 3 | 5 | 0 | 381 (9.3) | 4 (80) | 64 (16.8) |
|  | 2 | 1978 | 36 (1.8) | 198 | 1 | 1 | 200 | 8 | 534 (27.0) | 171 (86) | 14 (2.6) |
|  | 3 | 813 | 13 (1.6) | 7 | 3 | 0 | 10 | 0 | 60 (7.4) | 7 (70) | 3 (5) |
|  | 4 | 1942 | 6 (0.3) | 36 | 3 | 4 | 41 | 2 | 197 (10.1) | 36 (84) | 4 (2) |
|  | 5 | 1288 | 3 (0.2) | 1 | 0 | 3 | 4 | 0 | 91 (7.1) | 3 (75) | 0 (0) |
|  | 6 | 2450 | 10 (0.4) | 2 | 39 | 9 | 46 | 0 | 338 (13.8) | 27 (59) | 5 (1.5) |
|  | 7 | 2021 | 0 (0) | 1 | 0 | -<sup>e</sup> | 1 | 0 | 108 (5.3) | 1 (100) | 0 (0.9) |
|  | 8<sup>g</sup> | 366 | 4 (1.1) | 17 | 2 | 0 | 19 | 0 | 65 (17.8) | 19 (100) | 0 (0) |
|  | 9<sup>g</sup> | 1966 | 33 (1.1) | 3 | 5 | -<sup>e</sup> | 8 | 0 | 130 (6.6) | 3 (38) | 3 (2.3) |
| Elicited<sup>e</sup> | 1 | 5830 | 23 (0.4) | 0 | 0 | 1 | 1 | 0 | 41 (0.7) | 0 (0) | 3 (7.3) |
|  | 2 | 9646 | 39 (0.4) | 146 | 0 | 0 | 146 | 6 | 206 (2.1) | 65 (45) | 4 (1.9) |
|  | 3 | 5469 | 88 (1.6) | 4 | 0 | 3 | 7 | 0 | 63 (1.1) | 4 (57) | 2 (3.2) |
|  | 4 | 6757 | 48 (0.7) | 97 | 2 | 6 | 99 | 6 | 243 (3.6) | 72 (73) | 11 (4.5) |
|  | 5 | 3126 | 0 (0) | 0 | 0 | 1 | 1 | 0 | 22 (0.7) | 0 (0) | 0 (0) |
|  | 6 | 5809 | 2 (0.0) | 10 | 18 | 1 | 28 | 0 | 88 (1.5) | 8 (30) | 0 (0) |
|  | 7 | 3152 | 3 (0.1) | 2 | 0 | - | 2 | 0 | 30 (0.9) | 0 (0) | 0 (0) |
|  | 8 | 2821 | 24 (0.9) | 1 | 0 | 1 | 2 | 0 | 53 (1.9) | 1 (50) | 1 (1.9) |
|  | 9 | 2914 | 24 (0.8) | 2 | 1 | - | 3 | 0<sup>f</sup> | 42 (5.2) | 1 (33) | 0 (0) |

<sup>a</sup> Total unique T-cell clonotypes detected in skin prior to initiation of vaccination (Day 0)
<sup>b</sup> Total unique T-cell clonotypes only detected in skin after initiation of vaccination (Day 10 or later)
<sup>c</sup> % reflects the proportion of skin clonotypes detected in blood of all prevalent or elicited clonotypes at any lesion-area skin time point
<sup>d</sup> % reflects the proportion of skin clonotypes present in 4 or 5 out of 5 (or 3 of 3 in persons 8 and 9) biopsies after day 0
<sup>e</sup> these participants did not have biopsies performed at days 180 and 190
<sup>f</sup> in this person one dose 3 expander was detected in blood
<sup>g</sup> PBMC were not available at day 0 from this person

To further characterize prevalent clonotypes and their persistence in tissue over time, we compared their detection in serial biopsies. Prevalent and elicited clonotypes were identified to be resident if they were present in >3/5 or >2/3 biopsies (for those 2 persons who did not have biopsies performed after day 40) after day 0. A median of 9.3% of prevalent clonotypes were resident in tissue after day 0, compared to 1.3% of elicited clonotypes (p = 0.004) (**Fig. S2a, b, Table 1**). Of expanders, a median of 80% of prevalent clonotypes compared to a median of 33% elicited clonotypes were resident after vaccination (p = 0.004 by paired Wilcoxon). In all, prevalent clonotypes were more persistent in tissue than vaccine-elicited clonotypes.

We compared persistence and expansion over vaccine doses 2 and 3. **Fig. S2c** displays the clonotypes that expanded ≥6-fold from day 30 to day 40 pre and post dose 2 or were detected at >6 copies at day 40 after not being present prior to vaccination. P6 was the only participant in whom greater expansion was detected after dose 2 than dose 1. In all participants, 295 clonotypes increased in copy number after dose 2. Of those, 72 expanded ≥6 fold over the first dose and 11 expanded ≥6 fold over the second (day 30 to day 40); only one clonotype (in P6) expanded ≥6 fold over both doses. Over all participants, more expansion was detected after dose 1 than over the whole 6-month biopsy series or over the ten days before and after doses 2 and 3 (**Fig. 3d**, summary, **Table 1**).

### Disparity between skin and PBMC clonotypes

Next we aimed to quantify the representation of skin-based clonotypes and HSV-2-reactive CD4 *TRB* sequences from blood. *TRB* repertoire sequencing demonstrated that the T cell populations in the skin and blood shared few clonotypes. We analyzed the unique *TRB* sequences identified as clonotypes of interest in skin: 1) those expanding ≥6 fold after the first vaccine dose (n=266), 2) those appearing at ≥6 copies after the first vaccine dose (n=262), and 3) those that were detected in more than 70% of all lesion-area biopsies (n=1904 total, or 212 per participant on average) (**Table 1**). Across all samples, 0.7% of skin-based TRB sequences were detected among HSV-2-reactive CD4^+^ T cells from PBMCs (495 unique nucleotide sequences) which reflects 13% of PBMC-detected HSV-2-reactive CD4^+^ T cells (**Tables S3, S4**). The day 10 blood sample captured 4% of expanded (increased by ≥6 fold after dose 1) or highly elicited (detected at ≥6 copies at day 10) tissue-based clonotypes (22 of 528): 10 prevalent and 12 elicited clonotypes. It also captured 6.0% (114 of 1904) of clonotypes resident in skin, i.e., clonotypes detected in most skin biopsies after day 0 (either ≥4/5 or 3/3 for those persons with only 4 biopsies). The blood sample at day 0 did not capture any clonotypes expanded or elicited at ≥6 copies after vaccine dose 1 and captured only 7 (0.2%) of skin resident clonotypes (**Tables S3, S4**).

### Asymptomatic HSV shedding was common and unchanged over periods of observation

HSV-2 shedding (as defined by detection of HSV-2 DNA by PCR on a genital-area self-swab) during the trial was common, occurring in 5/9 participants during the first dose (from days 0-10), 3/9 during the second (from days 30-40), and 4/9 during the third (from days 180-190). All but 2 participants experienced shedding between dose 1 and 2. Persons with or without shedding were not observed to have alterations in their TRB repertoire or greater overlap with HSV-2-reactive CD4^+^ T cells from blood if shedding was situated prior to a biopsy (**Table S6**).

### Stability in V-J usage in skin and shifts in oligodominance after vaccination

To further investigate changes in *TRB* repertoires after vaccination, we evaluated the distribution of V-J usage within each participant’s *TRB* repertoire. In HSV-2 reactive CD4^+^ T cells from blood, V-J usage changed relatively dramatically between day 0 and day 10. By contrast, V-J usage in the arm was relatively constant across the vaccination series (**Fig. 4a, Fig. S3**). In genital area skin, vaccination resulted in a detectable shift of V-J usage in three of the nine participants after the first dose (P3, P4, and P5). In P3, BV24 and BV28 clonotypes were elicited and persisted through day 190. In P5, a BV18 clonotype was elicited at day 10 but was not detectable after day 30 (**Fig. 4a, Fig. S3)**. In P4, TRA/TRB sequencing of archived samples showed that prior to vaccination, a highly oligodominant V-J combination had been dominant for more than two years, and a novel oligodominant V-J combination became dominant at day 10. Prior to vaccination (day 0 and in 5 biopsies from two years before the vaccine trial) in P4, the most dominant V-J gene combination was TRAV21-01/TRAJ33-01 and TRBV07-09/TRBJ02-01. After vaccination, oligodominance switched to TRAV25-01/TRAJ15-01 and TRBV14-01/TRBJ02-05. **Fig. 4b** illustrates the oligoclonal pattern by TRAV/J and TRBV/J genes of the most prevalent TCR clonotypes two years prior to vaccination and at day 0 and day 10 after vaccination in genital area skin biopsies from P4. This vaccine-shifted, oligodominant V-J combination remained dominant in all subsequent biopsies, through day 190.

**Fig. 4.**
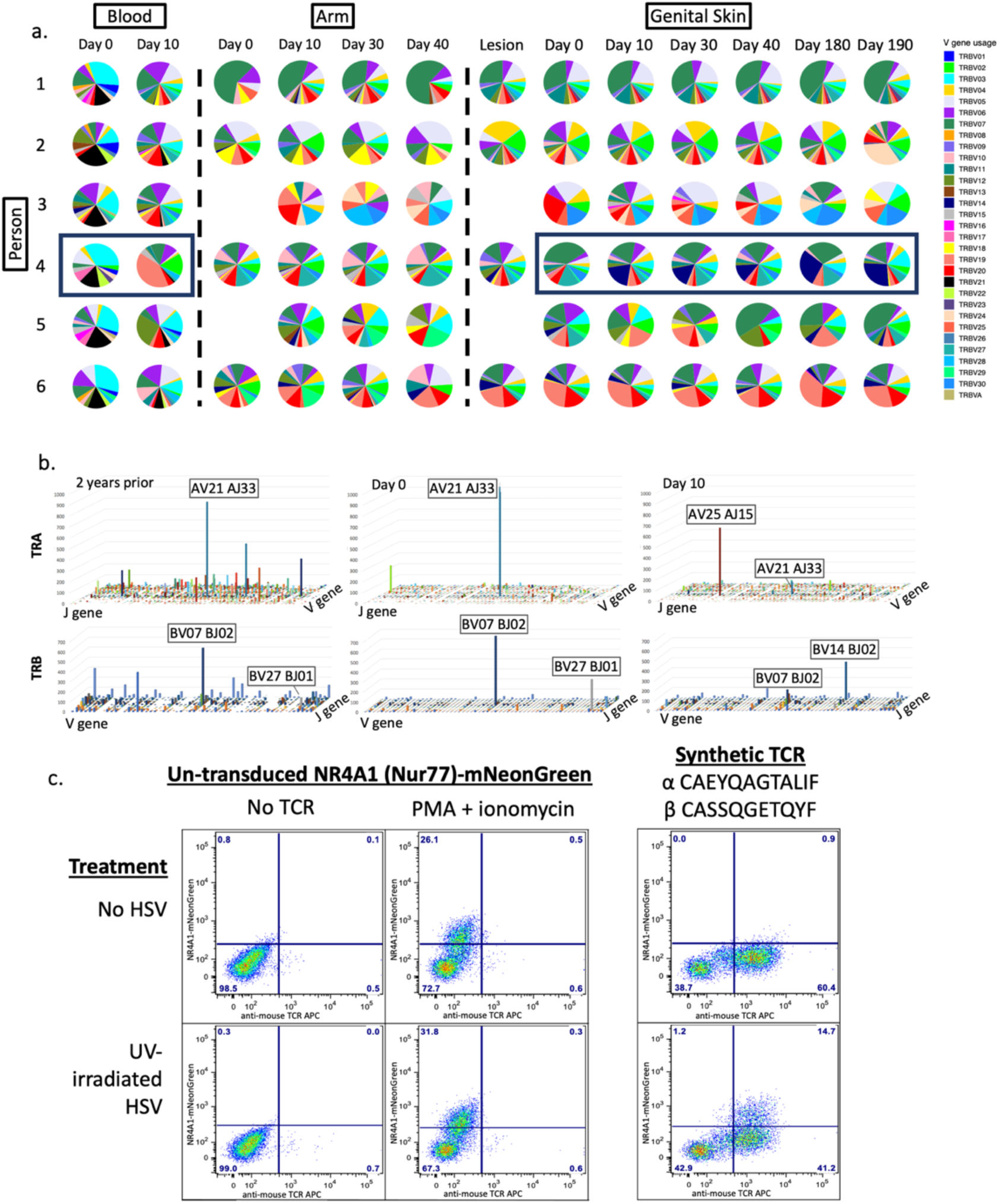
Evaluation of TRA/TRB V and J gene usage. (**a**) V gene usage in participants 1-6 across all sites, blood, arm, and genital skin, over time. Pie demonstrates proportion of the total repertoire represented by each gene. Skin site clonotypes were limited to those that were present at greater than 4 copies; all HSV-reactive CD4^+^ T cells from blood are shown. (**b**) Combination V and J gene usage in P4 showing the shift in immunodominance from two years prior to vaccination through ten days after the first vaccine dose. Z axis represents numbers of copies in each combination. X and y axes represent V and J genes listed sequentially. The V and J genes are labeled for the most abundant combinations. (**c**) HSV-2-specificity of a synthetic TCR composed of the most abundant TRA and TRB sequences from the day 10 sample in P4, using a Jurkat Nur77-mNeonGreen reporter cell system (right). Negative control (‘no TCR’, left) and positive control (PMA + Ionomycin, middle) for TCR stimulation and negative treatment (‘no HSV’, top) or experimental treatment (UV-irradiated HSV, bottom) to assess TCR specificity.

We next wanted to determine whether the most abundant post-vaccination TRA/TRB pair in P4 (TRAV25-01/TRAJ15-01 CAEYQAGTALIF; TRBV14-01/TRBJ02-05 CASSQGETQYF) was HSV-2-specific. Because it was not detected in blood at day 10 by AIM-based isolation of HSV-2-reactive T cells from PBMCs, a synthetic TCR with this TRA/TRB pair was expressed in a CD4-expressing Jurkat-based cell line with a NR4A1-mNeonGreen reporter system. NR4A1, also known as Nur77, is activated upon binding of the TCR to ligand. The transduced cell line showed reactivity in response to whole HSV-2 presented by autologous antigen presenting cells (APC), indicating HSV-2-specificity (**Fig 4c**). Specificity was refined by showing reactivity to a single HSV-2 protein, VP22, encoded by *UL49*, and a single VP22 peptide corresponding to residues 81-93 (ARPRRSASVAGSH) (**Fig. S4a, b**). To determine if this TCR had the characteristics of TCRs used by CD4^+^ T cells, we examined inhibition of activation of the reporter Jurkat line by HLA class II loci-specific blocking antibodies. A monoclonal antibody that blocks HLA-DP, but not those that block HLA-DR or DQ, was able to prevent reporter cell activation (**Fig. S4c**). Together with recognition of killed viral antigen, these data support that this TRA/TRB pair originated in a CD4^+^ T cell, indicating that vaccination elicited HSV-specific tissue-based CD4^+^ T cells not detected in PBMCs.

### Mucosal-associated invariant T (MAIT)-like TRA sequences are detectable in HSV lesion site skin but do not show expansion after vaccination

To discern the proportion of the skin-based TCR repertoire that may represent MAIT cells, as defined by TRA chains utilizing TRAV1-2 and TRAJ families 12, 20, or 33, we performed TRA sequencing of biopsies from P4 and P2. Overall, potential MAIT-like cells in both participants represented approximately 2% (0.9-2.6%) of the identified TRA sequences in HSV lesion site and control biopsies and did not expand or contract over time with vaccination (**Table 2**). Therefore, this vaccine does not appear to specifically modulate skin-based MAIT-like cells.

**Table 2.**
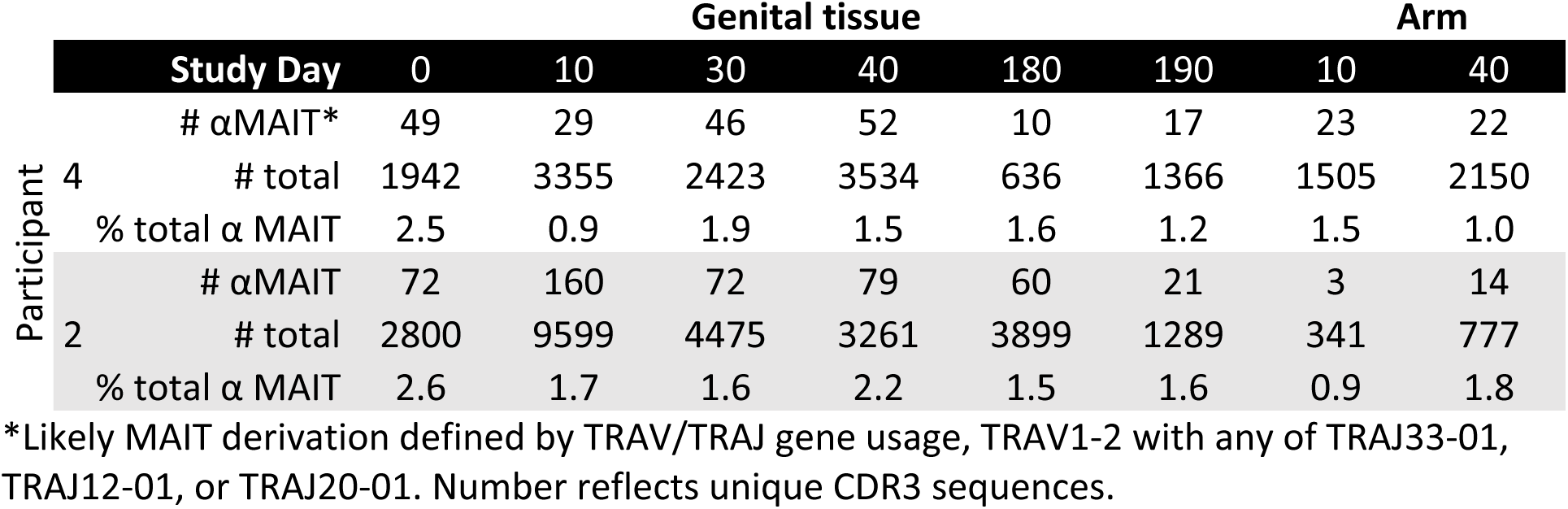
TRAV/TRAJ usage suggestive of MAIT cell derivation represent a small minority of the TCRα clonotypes detected in two participants over time.

### Mapping HSV-2 specificity of clonotypes detected in both PBMCs and skin biopsies

To explore the antigenic specificity of clonotypes that were shared in the blood and skin compartments in P4, we evaluated CD4^+^ T cells reactive to HSV-2 from day 10 PBMCs by single cell *TRA/TRB* sequencing and fine specificity determination (**Fig. S5**). HSV-2 reactivity was determined by expression of CD137 (4-1BB) by memory T cells after activation through TCR by UV-HSV186, yielding live, antigen-reactive cells for single-cell cloning directly ex vivo (*21*).

Among 192 clones generated from CD4^+^ T cells with high CD137 expression after stimulation with UV-HSV186, 190 proliferated in response to whole HSV-2 antigen, suggesting highly efficient enrichment of HSV-2 reactive clones (**Fig. S5a,b**). Paired *TRA/TRB* sequencing yielded a single productive *TRB* and either one or two productive *TRA* CDR3 sequences in 160 of these clones (**Fig. S5c, Table S7**). Of these 160, we selected 16 clones that had perfect *TRB* nucleotide sequence matches in genital skin and had characteristics of clonotypes of interest – either expanding ≥4 fold after the first dose or identified at 24 copies at day 10 – and determined their viral protein-level specificity using a library of all curated HSV-2 proteins (*33*). **Fig. 5a** outlines the number of copies detected in skin and blood and TCR specificity from all 16 clones. Reactivity was observed to proteins encoded by genes *ICP0, UL11, UL22, UL23, UL39*, *UL49* and *US6,* among others, indicating that among these overlapping clonotypes antigenic specificity did not seem to be related to their detection in both compartments. A representative example of HSV-2 protein UL11 specificity (clonotype 3 in **Fig. 5a**) is shown in **Fig. 5b**. This also confirmed HSV-2-specificity of 16 clonotypes in P4 that were observed to either expand from day 0 to day 10 or were newly detected in tissue after dose 1 at ≥4 copies, all of which were resident in tissue; clonotypes identified as clonotypes of interest. Therefore, in this participant vaccination induced broad CD4^+^ T cell reactivity in the skin at a site of previous HSV-2 reactivation.

**Fig. 5.**
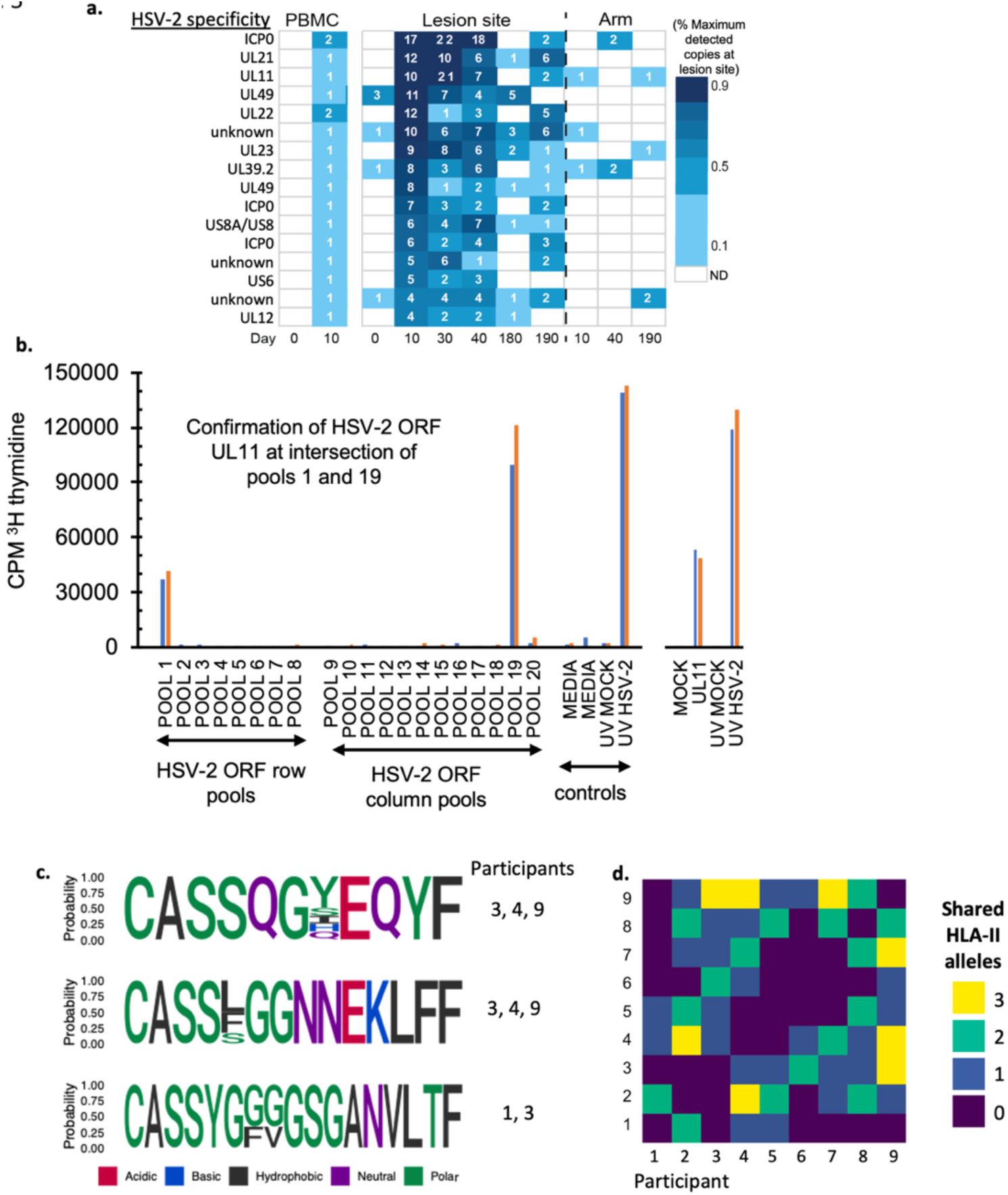
Representative data from fine specificity determination of blood CD4 T-cell clones overlapping with TCRβ CDR3 sequences detected in HSV lesion site biopsies. (**a**) Clonotype tracking with assigned specificity of 13 of the 16 clones queried in P4 by abundance and fold change in skin over dose one (day 0 to 10). (**b**) Example of fine-specificity mapping of a single clone confirmed to be specific to UL11. Both specimens were obtained from day 10 after HSV529 vaccination. At left is T-cell proliferation in response to matrix pools of HSV-2 antigens with positive and negative controls. Pools containing US11 (Pool 1, Pool 19) are positive. At right is confirmatory assay with recombinant UL11 and controls. Blue and orange bars represent replicate assays. (**c**) Logoplot representation of 3 sequence-similar (NN-distance <13) clonotype clusters in HSV-2 reactive CD4+ T cells from blood and clonotypes expanding after dose 1. (**d**) Number of shared HLA-II alleles (among DR, DQ, DP) between participants.

Analysis using a sequence-similarity algorithm (TCRdist3) was undertaken to determine the publicity of HSV-2-reactive clonotypes among these 9 participants with chronic HSV infection and their relationship with clonotypes observed to expand. Among all HSV-2-reactive or HSV-2-specific clonotypes from blood and 1100 total sequences observed to expand by or be elicited at ≥4 copies and using a conservative definition of TRB sequence similarity (<13 sequence-similarity distance units (nearest-neighbor (NN)-distance)), 38 clusters of clonotypes involving 3 or more clonotypes were identified including 10 with sharing between participants and 11 with sharing between site. Logoplots of 3 public clusters are shown in **Fig. 5c** and number of shared HLA-II alleles are shown in **Fig. 5d**.

## Discussion

Our study reveals several novel observations pertinent to immunotherapeutic vaccines or systemic immunotherapies for infectious, and likely also malignant, diseases. Immunization with a recombinant, replication-defective vaccine delivered intramuscularly in the arm elicited expansion of HSV-specific responses in blood and skin near a typical HSV lesion site in all participants. Demonstration of T cell expansion post vaccination was in many participants more sensitive through evaluation of skin biopsies than through PBMC sampling. While vaccination led to an increase in detection in blood of clonotypes also resident and highly abundant in skin, these represented a small minority of the total skin clonotypes and clonotypes of interest. *TRB* clonotypes that increased in abundance after vaccination in skin or appeared as vaccine-elicited in blood were often from the population of clonotypes present in skin prior to vaccination (both during quiescence at day 0 and/or in an active lesion at enrollment). Our findings are compatible with our current understanding of the immune response to chronic infection with HSV-2: there is a population of skin-resident T cells that are located at the site of reactivation and are poised to respond to viral challenge as it occurs (*28*, *30*, *34*). Moreover, among vaccine-elicited and vaccine-expanded clonotypes in skin, clonotypes already present at the time of vaccine initiation were more likely to remain resident in skin. Hence, for a sustained response, it is tissue-based clonotypes to which immunotherapy must be directed. Unfortunately, a viable and robust mechanism by which to pursue this in humans has not yet been identified. The vaccine we utilized was discontinued from further study due to the relatively poor immunogenicity based on standard assays, however the immune responses detected by TRB clonotype tracking were substantially different than predicted by PBMC ICS. Whether heightened tissue-based responses would be of clinical benefit, or whether tissue-based responses might predict clinical efficacy requires study. Novel technologies to recruit and bolster skin-based resident T cells should be a focus for the development of immunotherapeutic vaccines against HSV-2.

The uniqueness and persistence of the skin-resident population is shown most strikingly in P4, in whom TRB repertoire sequencing was possible for samples from two years prior to the present study. From these pre-trial biopsies, an oligoclonal population (>10% of TRB clonotypes derived from the same V-J combination) was detected in genital region biopsies and was not present in the arm or in HSV-reactive CD4^+^ T cells from PBMCs. A switch in the immunodominant oligoclonal “swarm” of clonotypes, from one dominant V-J combination to another, occurred after the first vaccine dose and was sustained for the full six months of this study with a single dominant *TRB*. By creation of a synthetic TCR, we confirmed that this vaccine-elicited, dominant T-cell clonotype originated from an HSV-2-reactive CD4^+^ T cell and its specificity was mapped to UL49.

Over all participants, vaccination, including repeated boosting doses, was not associated with an increase in the total cell population at the site of HSV-2 reactivation as measured by T-cell density, and indeed, boosting did not appear to enhance responses described here after dose 1 by T cell count or clonal tracking. In fact, the expansion response to subsequent doses appeared to diminish from dose 1 to doses 2 and 3. This cannot be explained by the available data presented here, but tissue-based study of subsequent vaccine trials may shed light on whether this may reflect antigen re-exposure fatigue or repertoire trimming. In all participants, selective increases in clonotypes were observed and detection of shared HSV-2-reactive TRB sequences increased after the first vaccine dose, albeit expansion varied considerably by participant. Determinants of such variability absolutely require further evaluation. These data indicate that immunogenicity studies of immunotherapeutic vaccines should carefully assess alterations in the TCR repertoire of the pathogen under study and not necessarily a total quantitative increase in antigen-specific T cells.

Within-person *TRB* nucleotide sequence identity with defined HSV-2 reactive clonotypes from PBMCs was used here as a surrogate indicator of HSV-2 specificity of tissue-based TRB sequences. This mechanism undoubtedly misses a large proportion of tissue-based HSV-2-specific T cells, such as the non-blood clonotype shown to be HSV-2-specific when expressed in a reporter system, but the true extent of HSV-2 specificity among tissue-resident T cells has not been defined. Alternative methods to expand the capacity to determine the specificity of tissue-based T cell clonotypes are in development, but clonal tracking is one novel method to expand the capacity to define T cell specificity into difficult to reach compartments.

The lack in overlap between skin and CD4^+^ T cells from PBMCs is magnified by the ability to select for CD4+ T cells with HSV-2 specificity from PBMC samples, which was not feasible for skin biopsies. Indeed, the contribution of CD8+ T cell clonotypes to the observed vaccine response in the skin is unknown, and although CD4+ T cell expansion was predicted by PBMC-based analysis in initial vaccine trials, this may not be representative of the response in skin. Altogether, even when selecting for those TCR clonotypes in greatest abundance in skin where overlap with PBMCs seems most likely, there was little representation in the HSV-2-enriched CD4+ T cell samples. In a chronic viral disease for which an effective vaccine has not been found after many years of trials, we wonder whether the focus on responses in PMBCs may partly explain previous failures. Our findings suggest that the inclusion of skin-based responses in the design of immunotherapeutic vaccines with a skin-based target may provide novel insights and perhaps bring an effective vaccine for HSV-2 and other chronic infections closer to reality. This study has shown conceptually that therapeutic targeting of T cells resident in genital skin sites by immunotherapeutic vaccination is possible.

## Materials and Methods

*Study design*: This study was designed as an open-label Phase 1 study to observe the shifts in the *TRB* repertoire in the skin at the site of HSV-2 reactivation in comparison to HSV-2-reactive CD4^+^ T cells obtained from peripheral blood and biopsies from an uninvolved arm site over the course of a 6-month, 3-dose vaccination study with replication-incompetent vaccine candidate HSV529 (Sanofi). This study was specifically designed to evaluate whether immunologic changes occurred in sequential genital skin biopsies following HSV529 vaccination. Arm biopsies, lesion-site biopsies during a symptomatic lesion, and lesion-site biopsies following HSV suppression with acyclovir were used in each participant as internal controls – all participants received vaccine (Study schema in **Table S1**, **Fig. 1a**). Demographic information is shown in **Table S2.**

*Participant recruitment and enrollment:* Healthy men and women with a history of symptomatic genital herpes in areas amenable to biopsy were recruited to the University of Washington Virology Research Clinic (UW-VRC) in Seattle, WA. Sex was not considered as a biological variable. HSV-2 seropositivity was confirmed by Western blot (*36*). Persons living with HIV were excluded.

*Vaccine:* The vaccine contained 0.5ml (1× 10^7^ pfu/dose) of HSV529, a replication-deficient, double-deletion (*UL5* and *UL29*) HSV-2 strain (Sanofi Pasteur)(*37*). Doses were delivered by intramuscular injection into the deltoid at days 0, 30 and 180. (**Fig. 1a, Table S1**).

*HSV lesion-area skin biopsies:* Genital-area skin biopsies (3mm) were obtained at the site of HSV-2 reactivation, either confirmed by observation of a lesion at a study visit or by reported history. At enrollment, participants had a baseline biopsy performed at the site of a lesion, if present, or at the site of the most frequent recurrence, and received four weeks of valacyclovir 500mg daily to suppress HSV-2 reactivation. Participants stopped valacyclovir 3 days prior to vaccination. Lesion site skin biopsies were performed prior to each vaccine dose on days 0, 30, and 180, and 10 days after each dose at days 10, 40, and 190 (*14*). Non-HSV-involved skin biopsies were obtained from the arm at days 0, 10, 40 and 190. Biopsy samples were fresh frozen in optimum cutting temperature (OCT) compound. Participants performed self-swabbing of the genital area daily for the first 30 study days and for 10 days after each vaccine dose. Detection of HSV DNA by PCR was performed as described (*38*).

*DNA extraction and sequencing:* A 1×3mm cross-section (inclusive of epidermis and dermis) from each biopsy was digested with proteinase-K and genomic DNA (gDNA) was extracted by spin column (Qiagen DNeasy). *TRB* repertoire sequencing was performed from gDNA samples by Adaptive Biotechnologies using the ImmunoSEQ assay (*39*). TCRα (*TRA*) chain sequencing (Adaptive Biotechnologies) from the same gDNA samples was performed for two participants. Adaptive TCR sequencing is a gDNA PCR-based platform in which the number of *TRB* sequence reads is proportional to the number of T cells bearing that unique sequence in each sample (*40*). In our experience, a variable but small proportion of HSV-2-specific T-cell clonotypes with the same *TRB* nucleotide sequence will be paired with different *TRA* sequences (*41*). The number of unique *TRB* sequences (including *CDR3*, V, D and J chain regions) is therefore a surrogate for the number of cells of that clonotype. *CDR3* nucleotide sequences plus V and J chain region identifications were used to compare persistence and detection of a specific clonotype within samples from a single participant. CDR3 amino acid sequences (with V and J chain usage) were used to compare clonotype publicity across multiple participants.

*Isolation of PBMCs*: In the initial trial of HSV529, the majority of alterations in HSV-2 specific T cell responses in PBMCs from HSV-2 seropositive participants were in the CD4^+^ T cell population (*14*). HSV-2-reactive CD4^+^ T cells were enriched from PBMCs obtained at days 0 and 10 by two methods. In the first, PBMCs were incubated with UV-inactivated cell-associated HSV-2 strain 186 (UV-HSV186) for 18 hours and stained for IFN-γ and IL-2 by intracellular staining (modified from Moss *et al.* (*20*)). Cytokine-producing cells were sorted, DNA was prepared with the Qiagen blood kit, and *TRB* sequencing performed as above (**Fig. S1**). In one participant (P4), at day 10 two selection procedures were performed iteratively to enhance the detection of HSV-2-reactive T cells from PBMCs. After incubation with UV-HSV186, CD3^+^CD4^+^CD8^-^CD137^high^ cells were sorted and expanded polyclonally (modified from Jing *et al.* (*21*)), then stimulated again with UV-HSV186 and stained for IFN-γ and IL-2 by intracellular staining (**Fig. S1**, method 2). gDNA extraction and *TRB* sequencing of bulk cytokine-producing cells was performed as above.

*Confirmation of HSV-2 specificity and determination of target epitope in PBMC-based clonotypes*: Live, CD3^+^CD4^+^CD137^+^ cells (n= 960) from frozen PBMC obtained from P4 at day 10 post-dose 1 were single-cell sorted and cloned as published (*17*). We selected 192 random clones for functional screening with autologous irradiated PBMC as antigen-presenting cells (APC), UV-HSV186 or UV-mock virus control as antigens, and tritiated thymidine incorporation proliferation assay to determine T cell proliferation in response to the given stimuli. We performed paired *TRA* and *TRB CDR3* sequencing on aliquots of ∼ 100 cells per clone using a SMARTSeq-2 cDNA procedure (*42*). The specificity of selected clones was determined to the level of the antigenic HSV-2 open reading frame (ORF) as previously described (*33*). Briefly, each HSV-2 ORF was cloned and expressed in vitro. T cell clones were expanded once as described (*18*). HSV-2 proteins were queried in proliferation assays as pools of 8 to 12 proteins/pool in matrix array at final concentrations of ∼1:5000 of each protein. To confirm specificity, HSV-2 protein(s) from pools at the intersection(s) of positive rows and columns were employed as antigens in subsequent proliferation assays.

*Development of reporter cell line:* To create the NR4A1_NeonGreen TCR reporter, the coding sequence of mNeonGreen was integrated in-frame into the *NR4A1* locus before the stop codon using CRISPR-induced homology directed repair (*43*). The mNeonGreen coding sequence (*44*) (IDT, Coralville IA) flanked by a 5’ 334bp homology arm, 5’ T2A element and a 315bp 3’ homology arm was electroporated with recombinant spCas9 (IDT) and *NR4A1* CRISPR guide RNA (IDT, sequence: AUGAAGAUCUUGUCAAUGAU) into Jurkat clone E6-1 cells (ATCC). Cells were cloned by limiting dilution and the clone with the highest signal upon PMA (5ng/ml) – ionomycin (5µg/ml) (Invivogen) stimulation was identified. The obtained reporter cell line was further modified by CRISPR knock out of *TRA*/*TRB* expression (gRNA sequences: AGAGUCUCUCAGCUGGUACA, CAAACACAGCGACCUUGGGU (IDT)); cells with successful knock out were sorted for lack of CD3 expression. The final NR4A1_NeonGreen TCR reporter showed upregulation of the reporter signal according to TCR signal strength, mimicking regulation of the endogenous *NR4A1* locus (*45*).

*Creation and HSV-2-specificity testing of synthetic TCR:* Full-length, codon-optimized, TCR genes were synthesized (IDT) with *TRB* preceding a P2A sequence. *TRA/TRB* pairing was promoted by partially replacing portions of the *TRA* and *TRB* constant regions with murine homologs with added cysteine residues. An epitope on the extracellular domain of murine *TRB* enabled flow cytometry monitoring of transduction as previously reported (*46*). The TCR inserts were cloned into pRRLSIN.cPPT.MSCV/GFP.WPRE (*47*) via Gibson assembly (New England Biolabs, Ipswich MA). After confirmation by sequencing, lentivirus was produced using a third-generation system (Addgene, Watertown MA) on LentiX-293T cells (Takada Bio, Mountain View CA)(*46*).

Sequences are available on GenBank (MZ821077-MZ821078). *<u>Specificity Assay</u>:* 200,000 synthetic TCR-transduced reporter cells were co-cultured with 200,000 participant-matched PBMC and 1/100 dilution of UV-HSV186 virus or mock for 16-20 hours. Un-transduced and PMA/Ionomycin-activated reporter cells were used as controls. Cells were labeled with Live Dead Aqua (Invitrogen) and anti-mouse TCR APC (BD Pharmingen) and analyzed on a FACS Canto II for expression of mNeongreen. *<u>Protein-level Specificity Assay</u>*: Protein-level specificity was assigned by measuring mNeongreen expression after 18-hour incubation of TCR-transduced reporter cells with matrix-pooled protein antigens (as above) spanning the HSV-2 proteome. Protein-level specificity was confirmed by a comparison between the HSV-2 protein of interest and a non-reacting protein at a range of concentrations (1:400 – 1:1600) for 18-24 hours. *<u>Peptide-level Specificity Assay:</u>* Peptide-level specificity was assigned by exposing the TCR-transduced reporter cells to pooled, linear peptides (1μg/ml) covering the UL49 sequence. *<u>Blocking Assay</u>:* Monoclonal antibodies to HLA-DP, DQ, and DR at a 1:40 final dilution were added to the specificity assays above in which UV-HSV186 was also serially diluted (1:100, 1:1000) to determine HLA specificity (*17*).

*Immunofluorescence staining*: To evaluate the effects of vaccination on T cell infiltration, we evaluated T cell spatial localization and skin density as previously described (*30*). Thawed, 8μm skin sections were washed, fixed in acetone, blocked with 1× casein (Vector Laboratories), 2% bovine serum albumin (BSA), and 5% normal human sera (NHS), and incubated for one hour in blocking solution at room temperature with mouse anti-human CD4 (1:500; Biolegend), and tagged with Alexa-Fluor 488 by tyramide signal amplification (TSA) (Invitrogen), followed by staining with Alexa-Fluor 647-labeled mouse anti–human CD8^+^ (1:100; Caltag Laboratories) overnight at 4°C. After nuclear staining with DAPI (Fluka), skin sections were mounted in Mowiol 40-88 containing 2.5% wt/vol DABCO (Sigma-Aldrich). Images were captured with Nikon Eclipse Ti with NIS-Elements AR Software (v4.40) and viewed in Fiji v2.9 (*48*). CD8^+^ and CD4^+^ T cells were counted manually (in triplicate) in fields of 650μm^2^, including the epidermis, dermal-epidermal junction and the upper dermis (*49*). To evaluate whether vaccination induced an upregulation of cell surface markers associated with tissue homing and residency, immunofluorescence with an antibody combination of mouse anti-human CD4 followed by TSA amplification, Alexa-Fluor 647 mouse anti-human CD69 (1:50; Biolegend) and rabbit anti-human CXCR3 (1:50, Abcam) with Alexa-Fluor 546 donkey anti-rabbit secondary (1:100, Invitrogen) was performed, as above.

*Computational and statistical analysis:* Comparison of T cell count and density in tissue as well as number of unique *TRB* sequences (clonotypes) between time points, cell populations, and blood/biopsy sites was performed by paired Wilcoxon signed-rank tests. For comparison of T cell count in tissue, the post-vaccination lesion area biopsies were tested against the arm control and the day 0 baseline biopsy specimens. For comparison of clonotype abundance, lesion area biopsy time points were compared against corresponding arm biopsy time points (10, 40, and 190 days after dose 1). To compute fold change in abundance over time when clonotypes were not present at both time points, a minimum abundance level of 1 copy was assigned to absent clonotypes. *TRA/TRB* sequence data was exported from the Adaptive Biotechnologies platform and analyzed offline. Analyses were performed using SAS 9.4 (SAS institute), Python 3.6.5 and R 3.4.1 (R Core Team). Non-productive sequences and those without V and J region identification were excluded from analysis. Statistical significance was defined as a two-sided p value ≤0.05. Sequence similarity was evaluated for unique CDR3 amino acid sequences from blood (limited to clonotypes observed at >1 copy from the sample that underwent ex vivo expansion prior to sequencing) and those expanding after vaccination in skin using TCRdist3 (v0.2.2)(*50*, *51*). Clonotypes considered to be similar were restricted to those within 13 nearest-neighbor (NN)-distance units, which would allow up to 4 exchanges of biochemically similar amino acids or 1 biochemically dissimilar amino acid or deletion in the CDR3 region, or up to 12 exchanges in other regions.) Logoplots were created with ggseqlogo.

*Study Approval:* Participants provided informed consent to participate in the trial. The University of Washington Human Subjects Division approved the study procedures. The trial was monitored by a Data Safety Monitoring Board. The trial was registered on clinicaltrials.gov, NCT02571166.

## Data Availability

The TCR sequencing data that support the findings of this study are available for review at Adaptive Biotechnologies, https://clients.adaptivebiotech.com with the following credentials: **Email**, **Password**: ford-2019-review. These data will be made public at publication. The custom code used for these analyses is available here or at reasonable request from the corresponding author: https://github.com/esford3/HSV529_manuscript

https://github.com/alvason/visualizing_Tcell_response_after_HSV2_vaccine

## Acknowledgements

The authors would like to express their appreciation to Amanda Woodward and Mindy Miner for their assistance in editing and revising the final manuscript, and to Khamsone Phasouk, Sijie Sun, Alexis Klock, and Lei Jin for their laboratory expertise.

## Author contributions

Study design: CJ, ASM, JZ, LC

TCR data analysis and bioinformatics: ESF, AL, LJ, ASM, DMK, LC

PBMC-based T cell assays and analysis: LD, LJ, KJL, KB, MO, DMK

Skin biopsy analysis: ESF, AK, JZ

Synthetic TCR specificity: KD, ME, AC

Participant recruitment and clinical guidance: CJ

Statistical analysis: ESF, ASM, LC

Consulting statistician: ASM

Vaccine acquisition and regulatory guidance: JT, SG, LC

Manuscript preparation: ESF, LC, KJL, DMK, CJ, ASM

All authors reviewed and approved of the manuscript.

## List of Supplementary Materials

**Fig. S1.** Experimental procedure to isolate HSV-reactive CD4+ T cells.

**Fig. S2.** All prevalent and elicited dose 1 and dose 2 expanding clonotypes from skin and cross-detection in blood and arm.

**Fig. S3.** TRBV and TRBJ gene usage in blood, arm, and genital skin.

**Fig. S4.** Determination of UL-49 reactivity and HLA-DP restriction of transgenic, vaccine-expanded immunodominant TCR from P4.

**Fig. S5.** Gating strategy and CMP results to identify HSV-specificity of clonotypes.

**Table S1.** Procedures and study visits.

**Table S2**. Demographic characteristics of participants enrolled in the vaccine and preliminary arms.

**Table S3.** Unique clonotypes identified as HSV-2-reactive in CD4+ T cells from blood and their detection in tissue.

**Table S4a.** All prevalent clonotypes detected in blood and skin.

**Table S4b.** All elicited clonotypes detected in blood and skin.

**Table S5a.** All prevalent clonotypes expanding over dose 1.

**Table S5b.** All elicited clonotypes expanding over dose 1.

**Table S5c.** All clonotypes expanding over dose 2.

**Table S6.** Detection of HSV-2 by genital-area swab (viral shedding) by study day range.

**Table S7.** All HSV-specific clonotypes from P4.

## Supplementary Methods

### Supplementary Fig. Legends

**Fig. S1. Schema of selection procedures for HSV-2-reactive CD4+ T cells from PBMCs.** Method 1 represents selection based on intracellular detection of IFN-γ or IL-2 after stimulation with UV-HSV186. Method 2 was performed in PBMCs from P4 at day 10 to enhance detection of HSV-2-reactive CD4^+^ T cells. After stimulation, CD137^high^ CD4^+^ T cells were expanded, then assayed for cytokine production as above, all cytokine-producing T cells were then submitted for TCRβ sequencing. Flow plots show example of ICS^+^ sorting in P2. Table inset shows the number of cells that were identified to be ICS^+^ and submitted for sequencing.

**Fig. S2. Clonal tracking of (a) prevalent and (b) elicited skin-based clonotypes ranked by fold change in abundance of copies from day 0 to day 10, or day 30 to 40 (c, d) in each participant.** Clonotypes from all participants were selected for and ranked by fold increase >6 from prior to vaccination to day 10 (prevalent clonotypes) or by increase over single copy to >6 copies at day 10 (elicited clonotypes). Each row represents a single clonotype (defined by nucleic acid sequence, the corresponding amino acid sequence is shown for readability), the number of copies are shown for each time point. Blank spots represent a time point where the clonotype was not detected. P1 had no elicited clonotypes at >6 copies, P5 had prevalent clonotypes that expanded at (but not greater than) 6-fold. (**c**) Prevalent and (**d**) elicited clonotypes are ranked by fold increase from day 30 to day 40. The number of sequences detected in PBMCs at day 10 in P4 was increased by the method of detection, where CD137-expressing clonotypes after UV-HSV186 exposure were expanded prior to selection by ICS and sequencing, these are denoted by $. Two participants (P7 and P9) did not have biopsies taken after day 40 and PBMCs from day 0 were not available in P6 and P8 (blocks without data are shaded in grey).

**Fig. S3. Relative use of V and J genes in TRB sequencing from HSV lesion site biopsies by participant and study day in comparison to those seen in HSV-2 reactive CD4+ T cells from PBMCs (“blood”) and arm biopsies**. V and J gene usage is displayed by proportion of the TRB sequences at each time point.

**Fig. S4. HSV-2 UL49-specificity of synthesized TCR from skin-derived TCRα/TCRβ sequencing of a clone observed to establish long-term immunodominance after dose 1 in P4. (a)** Protein-level specificity in the synthetic TCR from P4 and TCR-transduced reporter cell recognition (as measured by expression of mNeonGreen) of HSV-2 UL49 in an antigen concentration-dependent manner in comparison to lack of recognition of UL46. Gating is on live, lymphocyte subset. **(b)** Peptide-level specificity identified by screening of HSV-2 UL49 peptide pools, shown is percentage of cells co-expressing APC+/mNeonGreen+. **(c)** Confirmation of HLA-II use and thus CD4 origination of the TCR by blocking assay using monoclonal antibodies against HLA-DP, DQ, and DR. The TCR-transduced reporter cell line was stimulated with HSV2 strain 186 at final assay dilution of 1:100, 1:1000, or a mock exposure, in the presence of HLA-DP, DR, and DQ blocking antibodies at a final concentration of 1:40, with abrogation of the Nur77-mNeonGreen activation signal in the presence of an HLA-DP monoclonal antibody.

**Fig. S5. Identification of HSV-2-reactive CD4+ T cells from frozen PBMCs from P4 for determination of fine specificity, confirmation of reactivity and bulk sequencing results. (a)** Gating scheme for sorting of PBMC from P4, day 10 after first dose of HSV529 vaccine. After UV-HSV186 stimulation for 18 hours, brisk up-regulation of surface CD137 was detected and single cells were sorted and clonally expanded. **(b)** Lymphoproliferation results for 192 randomly selected clones presented as a histogram of net CPM for singlicate split-well analyses with whole UV-HSV186 antigen and mock antigen. **(c)** Summary of results of TCR CDR3 sequencing for the 192 CD4+ T-cell clones.

### Supplementary methods

Other inclusion criteria included age 18-55 years, willingness to use contraception for the duration of the study, and willingness to participate. Exclusion criteria included HIV-1 or HCV seropositivity, pregnancy or breast-feeding, BMI >35, serious chronic illness or immunocompromise (including use of immunomodulators).

## References and Notes

1. K. J. Looker, A. S. Magaret, K. M. E. Turner, P. Vickerman, S. L. Gottlieb, L. M. Newman, Global estimates of prevalent and incident herpes simplex virus type 2 infections in 2012. PLoS One. 10, 1–23 (2015).

2. K. J. Looker, J. A. R. Elmes, S. L. Gottlieb, J. T. Schiffer, P. Vickerman, K. M. E. Turner, M.-C. Boily, Effect of HSV-2 infection on subsequent HIV acquisition: An updated systematic review and meta-analysis. Lancet Infect. Dis. 3099, 1–14 (2017).

3. K. J. Looker, A. S. Magaret, M. T. May, K. M. E. Turner, P. Vickerman, L. M. Newman, S. L. Gottlieb, First estimates of the global and regional incidence of neonatal herpes infection. Lancet Glob. Heal. 5, 1–10 (2017).

4. C. M. Posavad, D. M. Koelle, M. F. Shaughnessy, L. Corey, Severe genital herpes infections in HIV-infected individuals with impaired herpes simplex virus-specific CD8+ cytotoxic T lymphocyte responses. Proc. Natl. Acad. Sci. U. S. A. 94, 10289–94 (1997).

5. D. I. Bernstein, A. Wald, T. Warren, K. Fife, S. Tyring, P. Lee, N. Van Wagoner, A. Magaret, J. B. Flechtner, S. Tasker, J. Chan, A. Morris, S. Hetherington, Therapeutic vaccine for genital herpes simplex virus-2 infection: Findings from a randomized trial. J. Infect. Dis. 215, 856–864 (2017).

6. N. Van Wagoner, W. Koltun, G. Lucksinger, T. Warren, S. Tyring, D. Bernstein, P. Leone, L. Panther, J. Lalezari, K. Fife, R. Novak, R. Beigi, J. Kriesel, J. Chan, S. Tasker, S. Hetherington, A. Wald, GEN-003 phase 2 interim results: Therapeutic vaccine for genital herpes significantly reduces viral shedding and genital lesions. Open Forum Infect. Dis. 2, 898 (2015).

7. A. Wald, T. Warren, K. Fife, S. Tyring, P. Lee, N. Van Wagoner, D. Bernstein, J. B. Flechtner, A. Magaret, A. Morris, S. Hetherington, Therapeutic HSV-2 vaccine (GEN003) results in durable reduction in genital lesions at 1 year. Open Forum Infect. Dis. 1, S55– S56 (2014).

8. S. E. Straus, L. Corey, R. L. Burke, B. Savarese, G. Barnum, P. R. Krause, R. G. Kost, J. L. Meier, R. Sekulovich, S. F. Adair, C. L. Dekker, Placebo-controlled trial of vaccination with recombinant glycoprotein D of herpes simplex virus type 2 for immunotherapy of genital herpes. Lancet. 343, 1460–1463 (1994).

9. M. Boukhvalova, J. McKay, A. Mbaye, H. Sanford-Crane, J. C. G. Blanco, A. Huber, B. C. Herold, Efficacy of the herpes simplex virus 2 (HSV-2) glycoprotein D/AS04 vaccine against genital HSV-2 and HSV-1 infection and disease in the cotton rat Sigmodon hispidus model. J. Virol. 89, 9825–9840 (2015).

10. R. B. Belshe, T. C. Heineman, D. I. Bernstein, A. R. Bellamy, M. Ewell, R. Van Der Most, C. D. Deal, Correlate of immune protection against HSV-1 genital disease in vaccinated women. J. Infect. Dis. 209, 828–836 (2014).

11. S. E. Straus, A. Wald, R. G. Kost, R. McKenzie, A. G. M. Langenberg, P. Hohman, J. Lekstrom, E. Cox, M. Nakamura, R. Sekulovich, A. Izu, C. L. Dekker, L. Corey, Immunotherapy of recurrent genital herpes with recombinant herpes simplex virus type 2 glycoproteins D and B: results of a placebo-controlled vaccine trial. J. Infect. Dis. 176, 1129–34 (1997).

12. D. M. Koelle, A. Magaret, C. L. McClurkan, M. L. Remington, T. Warren, F. Teofilovici, A. Wald, Phase I dose-escalation study of a monovalent heat shock protein 70-herpes simplex virus type 2 (HSV-2) peptide-based vaccine designed to prime or boost CD8 T-cell responses in HSV-naive and HSV-2-infected subjects. Clin. Vaccine Immunol. 15, 773–782 (2008).

13. A. Wald, D. M. Koelle, K. Fife, T. Warren, K. LeClair, R. M. Chicz, S. Monks, D. L. Levey, C. Musselli, P. K. Srivastava, Safety and immunogenicity of long HSV-2 peptides complexed with rhHsc70 in HSV-2 seropositive persons. Vaccine. 29, 8520–8529 (2011).

14. L. K. Dropulic, M. C. Oestreich, H. L. Pietz, K. J. Laing, S. Hunsberger, K. Lumbard, D. Garabedian, S. P. Turk, A. Chen, R. L. Hornung, C. Seshadri, M. T. Smith, N. A. Hosken, S. Phogat, L.-J. Chang, D. M. Koelle, K. Wang, J. I. Cohen, A randomized, double-blind, placebo-controlled, phase 1 study of a replication-defective herpes simplex virus 2 vaccine, HSV529, in adults with or without HSV infection. J. Infect. Dis. 20892, 1–11 (2019).

15. W. J. D. Ouwendijk, K. J. Laing, G. M. G. M. Verjans, D. M. Koelle, T-cell immunity to human alphaherpesviruses. Curr. Opin. Virol. 3, 452–460 (2013).

16. K. J. Laing, A. S. Magaret, D. E. Mueller, L. Zhao, C. Johnston, S. C. De Rosa, D. M. Koelle, A. Wald, L. Corey, Diversity in CD8+ T cell function and epitope breadth among persons with genital herpes. J. Clin. Immunol. (2010), doi:10.1007/s10875-010-9441-2.

17. D. M. Koelle, H. Abbo, A. Peck, K. Ziegweid, L. Corey, S. The, I. Diseases, N. May, Y. M. Spv-l, B. Nisperos, F. Hutchinson, Direct recovery of herpes simplex virus (HSV) –specific T lymphocyte clones from recurrent genital HSV-2 lesions. J. Infect. Dis. 169, 956–961 (1994).

18. D. M. Koelle, Expression cloning for the discovery of viral antigens and epitopes recognized by T cells. Methods. 29, 213–226 (2003).

19. L. Jing, J. T. Schiffer, T. M. Chong, J. J. Bruckner, D. H. Davies, P. L. Felgner, J. Haas, A. Wald, G. M. G. M. Verjans, D. M. Koelle, CD4 T-cell memory responses to viral infections of humans show pronounced immunodominance independent of duration or viral persistence. J. Virol. 87, 2617–27 (2013).

20. N. J. Moss, A. Magaret, K. J. Laing, A. S. Kask, M. Wang, K. E. Mark, J. T. Schiffer, A. Wald, D. M. Koelle, Peripheral blood CD4 T-cell and plasmacytoid dendritic cell (pDC) Reactivity to herpes simplex virus 2 and pDC number do not correlate with the clinical or virologic severity of recurrent genital herpes. J. Virol. 86, 9952–9963 (2012).

21. L. Jing, J. Haas, T. M. Chong, J. J. Bruckner, G. C. Dann, L. Dong, J. O. Marshak, C. L. McClurkan, T. N. Yamamoto, S. M. Bailer, K. J. Laing, A. Wald, G. M. G. M. Verjans, D. M. Koelle, Cross-presentation and genome-wide screening reveal candidate T cells antigens for a herpes simplex virus type 1 vaccine. J. Clin. Invest. 122, 654–673 (2012).

22. A. Sato, A. Suwanto, M. Okabe, S. Sato, T. Nochi, T. Imai, N. Koyanagi, J. Kunisawa, Y. Kawaguchi, H. Kiyono, Vaginal memory T cells induced by intranasal vaccination are critical for protective T cell recruitment and prevention of genital HSV-2 disease. J. Virol. 88, 13699–708 (2014).

23. J. T. Schiffer, L. Abu-Raddad, K. E. Mark, J. Zhu, S. Selke, D. M. Koelle, A. Wald, L. Corey, Mucosal host immune response predicts the severity and duration of herpes simplex virus-2 genital tract shedding episodes. Proc. Natl. Acad. Sci. U. S. A. 107, 18973–18978 (2010).

24. S. Ariotti, M. A. Hogenbirk, F. E. Dijkgraaf, L. L. Visser, M. E. Hoekstra, J.-Y. Song, H. Jacobs, J. B. Haanen, T. N. Schumacher, Skin-resident memory CD8^+^ T cells trigger a state of tissue-wide pathogen alert. Science. 346, 101–5 (2014).

25. N. Iijima, A. Iwasaki, A local macrophage chemokine network sustains protective tissue-resident memory CD4 T cells. Science. 346, 93–98 (2014).

26. H. Shin, A. Iwasaki, A vaccine strategy that protects against genital herpes by establishing local memory T cells. Nature. 491, 463–467 (2012).

27. T. Gebhardt, L. Wakim, L. Eidsmo, P. Reading, W. Heath, F. Carbone, Memory T cells in nonlymphoid tissue that provide enhanced local immunity during infection with herpes simplex virus. Nat. Immunol. 10, 524–530 (2009).

28. J. Zhu, T. Peng, C. Johnston, K. Phasouk, A. S. Kask, A. Klock, L. Jin, K. Diem, D. M. Koelle, A. Wald, H. Robins, L. Corey, Immune surveillance by CD8αα+ skin-resident T cells in human herpes virus infection. Nature. 497, 494–497 (2013).

29. T. Gebhardt, P. G. Whitney, A. Zaid, L. K. MacKay, A. G. Brooks, W. R. Heath, F. R. Carbone, S. N. Mueller, Different patterns of peripheral migration by memory CD4+ and CD8+ T cells. Nature. 477, 216–219 (2011).

30. J. Zhu, D. M. Koelle, J. Cao, J. Vazquez, M.-L. Huang, F. Hladik, A. Wald, L. Corey, Virus-specific CD8+ T cells accumulate near sensory nerve endings in genital skin during subclinical HSV-2 reactivation. J. Exp. Med. 204, 595–603 (2007).

31. J. Zhu, F. Hladik, A. Woodward, A. Klock, T. Peng, C. Johnston, M. Remington, A. Magaret, D. M. Koelle, A. Wald, L. Corey, Persistence of HIV-1 receptor-positive cells after HSV-2 reactivation is a potential mechanism for increased HIV-1 acquisition. Nat. Med. 15, 886– 892 (2009).

32. D. G. Coffey, LymphoSeq: Analyze high-throughput sequencing of T and B cell receptors. R Package version 1.18.0. (2020).

33. C. Johnston, J. Zhu, L. Jing, K. J. Laing, C. M. McClurkan, A. Klock, K. Diem, L. Jin, J. Stanaway, E. Tronstein, W. W. Kwok, M.-L. Huang, S. Selke, Y. Fong, A. Magaret, D. M. Koelle, A. Wald, L. Corey, Virologic and immunologic evidence of multifocal genital herpes simplex virus 2 infection. J. Virol. 88, 4921–31 (2014).

34. T. Peng, J. Zhu, K. Phasouk, D. M. Koelle, A. Wald, L. Corey, An effector phenotype of CD8+ T cells at the junction epithelium during clinical quiescence of herpes simplex virus 2 infection. J. Virol. 86, 10587–10596 (2012).

35. J. B. Flechtner, D. Long, S. Larson, V. Clemens, A. Baccari, L. Kien, J. Chan, M. Skoberne, M. Brudner, S. Hetherington, Immune responses elicited by the GEN-003 candidate HSV-2 therapeutic vaccine in a randomized controlled dose-ranging phase 1/2a trial. Vaccine. 34, 5314–5320 (2016).

36. R. L. Ashley, J. Militoni, F. Lee, A. Nahmias, L. Corey, Comparison of Western blot (immunoblot) and glycoprotein G-specific immunodot enzyme assay for detecting antibodies to herpes simplex virus types 1 and 2 in human sera. J. Clin. Microbiol. 26, 662–667 (1988).

37. X. Da Costa, M. F. Kramer, J. Zhu, M. A. Brockman, D. M. Knipe, Construction, phenotypic analysis, and immunogenicity of a UL5/UL29 double deletion mutant of herpes simplex virus 2. J. Virol. 74, 7963–7971 (2000).

38. K. E. Mark, A. Wald, A. S. Magaret, S. Selke, S. Kuntz, M. L. Huang, L. Corey, Rapidly cleared episodes of oral and anogenital herpes simplex virus shedding in HIV-infected adults. J. Acquir. Immune Defic. Syndr. 54, 482–488 (2010).

39. H. S. Robins, P. V Campregher, S. K. Srivastava, A. Wacher, C. J. Turtle, O. Kahsai, S. R. Riddell, E. H. Warren, C. S. Carlson, Comprehensive assessment of T-cell receptor beta-chain diversity in alpha-beta T cells. Blood. 114, 4099–107 (2009).

40. C. S. Carlson, R. O. Emerson, A. M. Sherwood, C. Desmarais, M. W. Chung, J. M. Parsons, M. S. Steen, M. A. LaMadrid-Herrmannsfeldt, D. W. Williamson, R. J. Livingston, D. Wu, B. L. Wood, M. J. Rieder, H. Robins, Using synthetic templates to design an unbiased multiplex PCR assay. Nat. Commun. 4, 1–9 (2013).

41. L. Dong, P. Li, T. Oenema, C. L. McClurkan, D. M. Koelle, Public TCR use by herpes simplex virus-2-specific human CD8 CTLs. J. Immunol. 184, 3063–3071 (2010).

42. L. Jing, M. Ott, C. D. Church, R. M. Kulikauskas, D. Ibrani, J. G. Iyer, O. K. Afanasiev, A. Colunga, M. M. Cook, H. Xie, A. L. Greninger, K. G. Paulson, A. G. Chapuis, S. Bhatia, P. Nghiem, D. M. Koelle, Prevalent and diverse intratumoral oncoprotein-specific CD8+ T cells within polyomavirus-driven merkel cell carcinomas. Cancer Immunol. Res. 8, 648– 659 (2020).

43. D. N. Nguyen, T. L. Roth, P. J. Li, P. A. Chen, R. Apathy, M. R. Mamedov, L. T. Vo, V. R. Tobin, D. Goodman, E. Shifrut, J. A. Bluestone, J. M. Puck, F. C. Szoka, A. Marson, Polymer-stabilized Cas9 nanoparticles and modified repair templates increase genome editing efficiency. Nat. Biotechnol. 38, 44–49 (2020).

44. N. C. Shaner, G. G. Lambert, A. Chammas, Y. Ni, P. J. Cranfill, M. A. Baird, B. R. Sell, J. R. Allen, R. N. Day, M. Israelsson, M. W. Davidson, J. Wang, A bright monomeric green fluorescent protein derived from Branchiostoma lanceolatum. Nat. Methods. 10, 407– 409 (2013).

45. J. F. Ashouri, A. Weiss, Endogenous Nur77 Is a specific indicator of antigen receptor signaling in human T and B cells. J. Immunol. 198, 657–668 (2017).

46. L. Jing, K. J. Laing, L. Dong, R. M. Russell, R. S. Barlow, J. G. Haas, M. S. Ramchandani, C. Johnston, S. Buus, A. J. Redwood, K. D. White, S. A. Mallal, E. J. Phillips, C. M. Posavad, A. Wald, D. M. Koelle, Extensive CD4 and CD8 T cell cross-reactivity between alphaherpesviruses. J. Immunol. 196, 2205–2218 (2016).

47. T. M. Schmitt, D. H. Aggen, K. Ishida-Tsubota, S. Ochsenreither, D. M. Kranz, P. D. Greenberg, Generation of TCRs of higher affinity by antigen-driven differentiation of progenitor T cells in vitro. Nat. Biotechnol. 35, 1188–1195 (2017).

48. J. Schindelin, I. Arganda-Carreras, E. Frise, V. Kaynig, M. Longair, T. Pietzsch, S. Preibisch, C. Rueden, S. Saalfeld, B. Schmid, J.-Y. Tinevez, D. J. White, V. Hartenstein, K. Eliceiri, P. Tomancak, A. Cardona, Fiji: An open-source platform for biological-image analysis. Nat. Methods. 9, 676–682 (2012).

49. K. Diem, A. Magaret, A. Klock, L. Jin, J. Zhu, L. Corey, Image analysis for accurately counting CD4+ and CD8+ T cells in human tissue. J. Virol. Methods. 222, 117–121 (2015).

50. K. Mayer-Blackwell, S. Schattgen, L. Cohen-Lavi, J. C. Crawford, A. Souquette, J. A. Gaevert, T. Hertz, P. G. Thomas, P. G. Bradley, A. Fiore-Gartland, TCR meta-clonotypes for biomarker discovery with tcrdist3 enabled identification of public, HLA-restricted clusters of SARS-CoV-2 TCRs. Elife. 10, 1–32 (2021).

51. P. Dash, A. J. Fiore-Gartland, T. Hertz, G. C. Wang, S. Sharma, A. Souquette, J. C. Crawford, E. B. Clemens, T. H. O. Nguyen, K. Kedzierska, N. L. La Gruta, P. Bradley, P. G. Thomas, Quantifiable predictive features define epitope-specific T cell receptor repertoires. Nature. 547, 89–93 (2017).

