## Supplemental Figures for "Expansion of the HSV-2-specific T cell repertoire in skin after immunotherapeutic HSV-2 vaccine"

### Supplementary figures and tables

12.30.23

**Supplementary Figures:**
**Supplementary Figure 1.** Experimental procedure to isolate HSV-reactive CD4+ T cells
**Supplementary Figure 2.** All prevalent and elicited dose 1 and dose 2 expanding clonotypes from skin and cross-detection in blood and arm.
**Supplementary Figure 3.** TRBV and TRBJ gene usage in blood, arm, and genital skin
**Supplementary Figure 4.** Determination of UL-49 reactivity and HLA-DP restriction of transgenic, vaccine-expanded immunodominant TCR from participant 4
**Supplementary Figure 5.** Gating strategy and CMP results to identify HSV-specificity of clonotypes

**Supplementary Tables:**
**Supplementary Table 1.** Procedures and study visits
**Supplementary Table 2**. Demographic characteristics of participants enrolled in the vaccine and preliminary arms.
**Supplementary Table 3.** Summary of unique clonotypes identified as HSV-2-reactive in CD4+ T cells from blood and their detection in tissue.
**Supplementary Table 4a.** All prevalent clonotypes detected in blood and skin.
**Supplementary Table 4b.** All elicited clonotypes detected in blood and skin.
**Supplementary Table 5a.** All prevalent clonotypes expanding over dose 1.
**Supplementary Table 5b.** All elicited clonotypes expanding over dose 1.
**Supplementary Table 5c.** All clonotypes expanding over dose 2.
**Supplementary Table 6.** Detection of HSV-2 by genital-area swab (viral shedding) by study day range.
**Supplementary Table 7.** All HSV-specific clonotypes from participant 4.

Supplementary Figure 1.

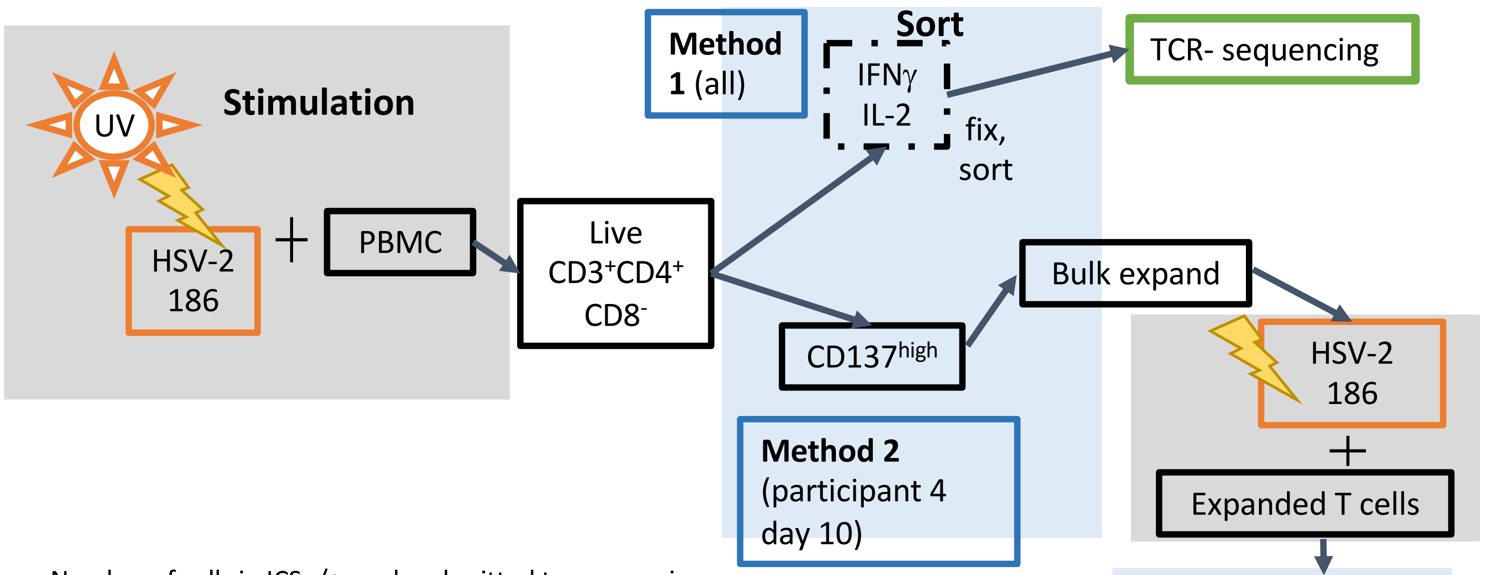


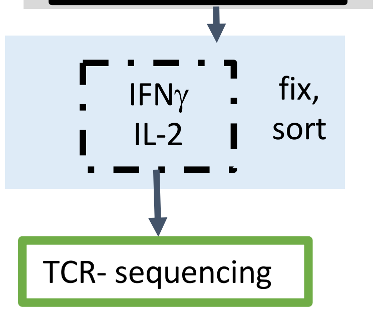


Number of cells in ICS -/+ pools submitted to sequencing

| Participant | Day 0 – neg pool | Day 0 submitted | Day 10 – neg pool | Day 10 submitted |
|---|---|---|---|---|
| 1 | 289082 | 642 | 401533 | 2063 |
| 2 | 120855 | 768 | 360478 | 1478 |
| 3 | 386782 | 1032 | 440487 | 9688 |
| 4 | 297566 | 7696 | | |
| 5 | 264471 | 626 | 95977 | 430 |
| 6 | 142644 | 165 | 107899 | 403 |
| 7 | 238454 | 125 | 124473 | 343 |
| 8 | NA | NA | 451953 | 13161 |
| 9 | NA | NA | 314766 | 10686 |

Example – Method 1
ICS fix/sort, Participant 2

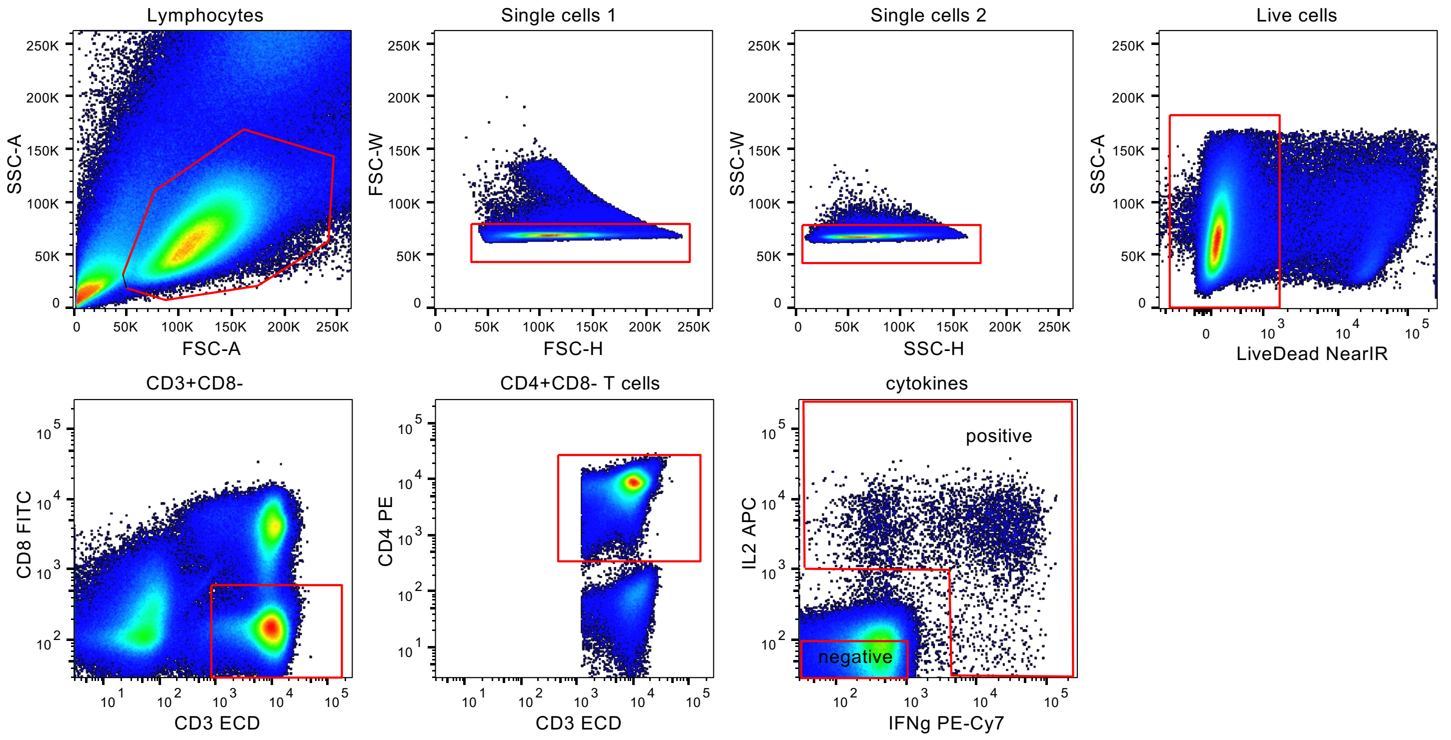

**Supplementary Figure 2.** All prevalent and elicited clonotypes expanding over dose 1

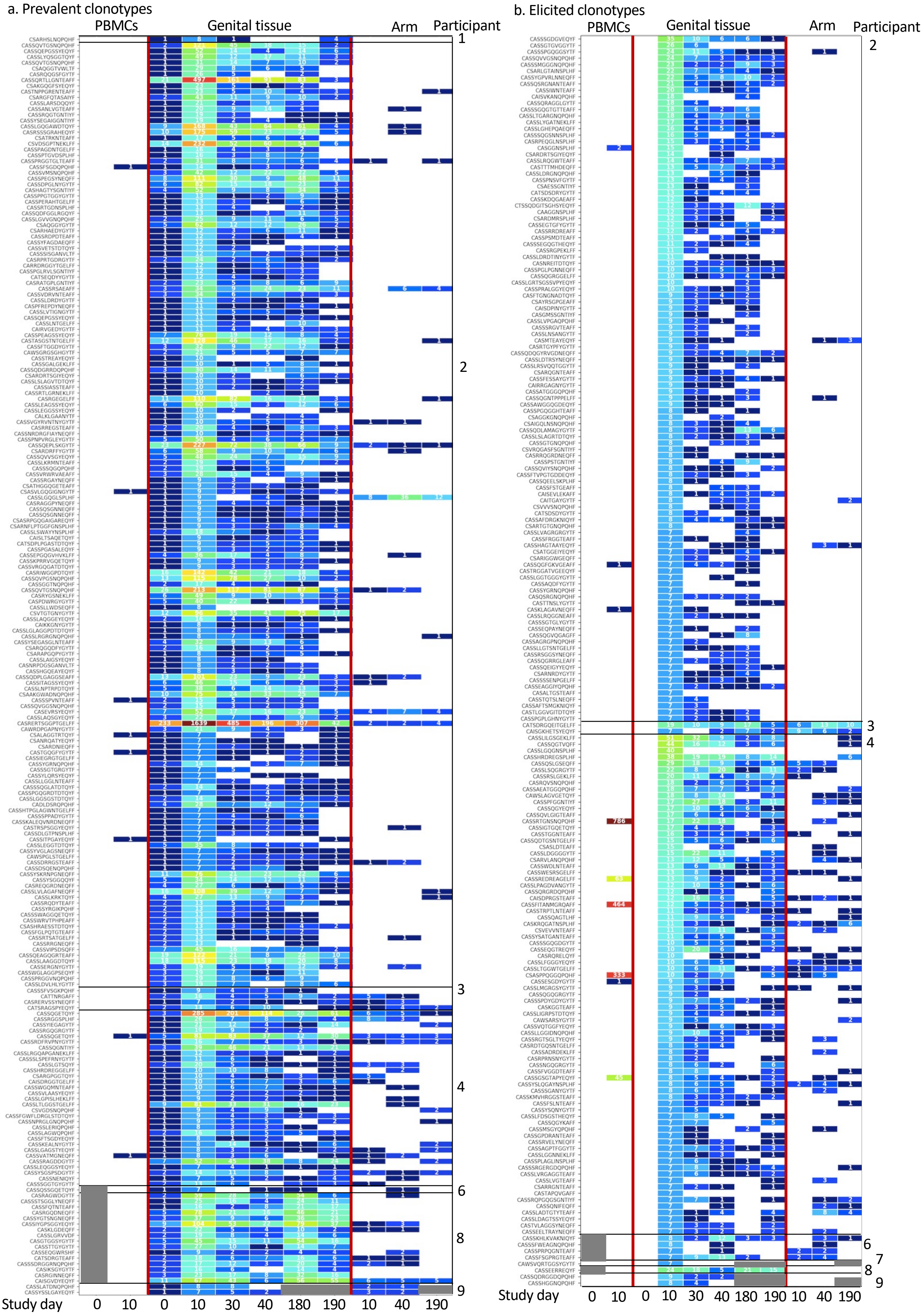

**Supplementary Figure 2.** All prevalent and elicited clonotypes expanding over dose 2

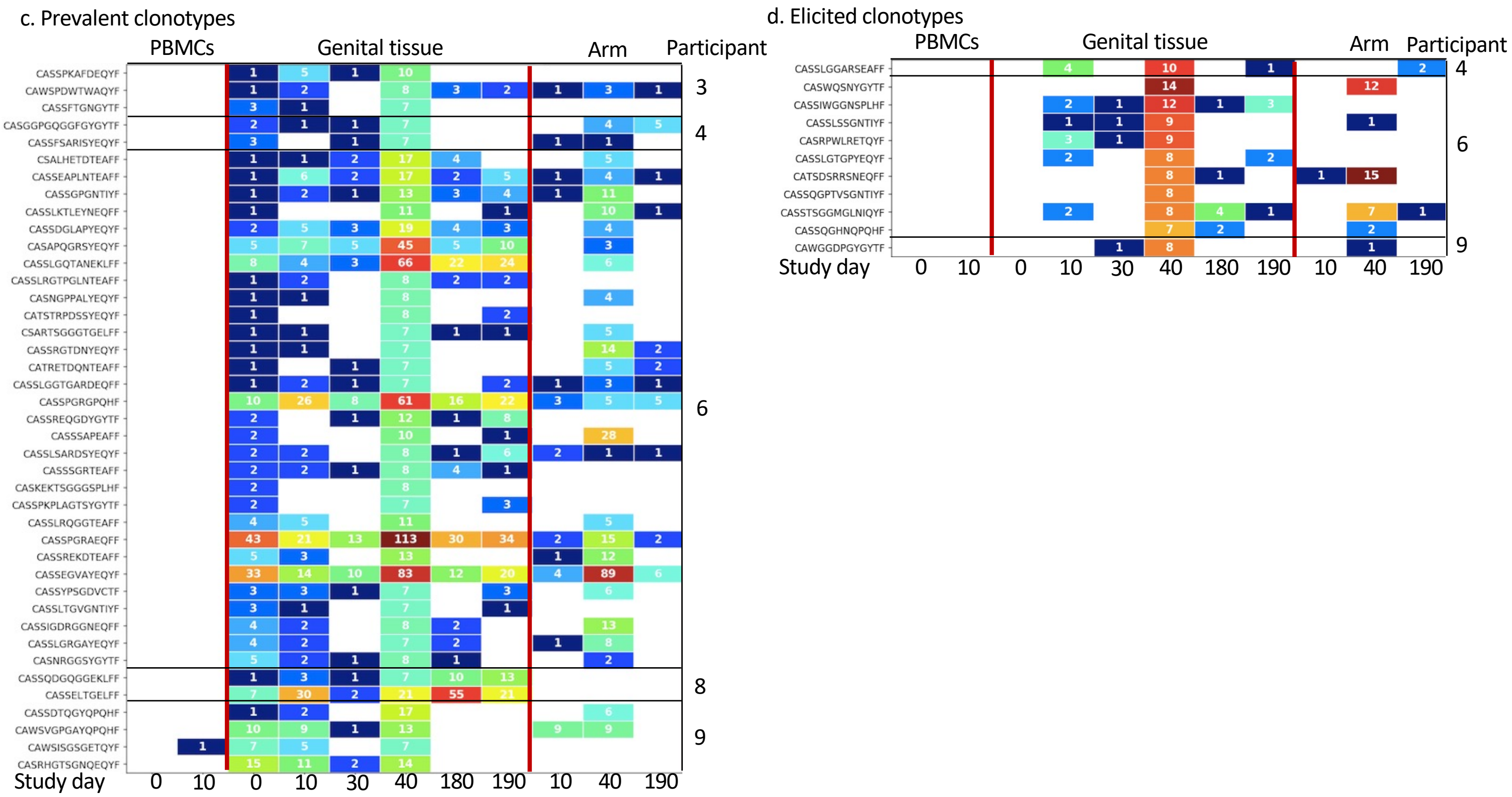

**Supplementary Figure 3** – TRBV and TRBJ usage in all participants

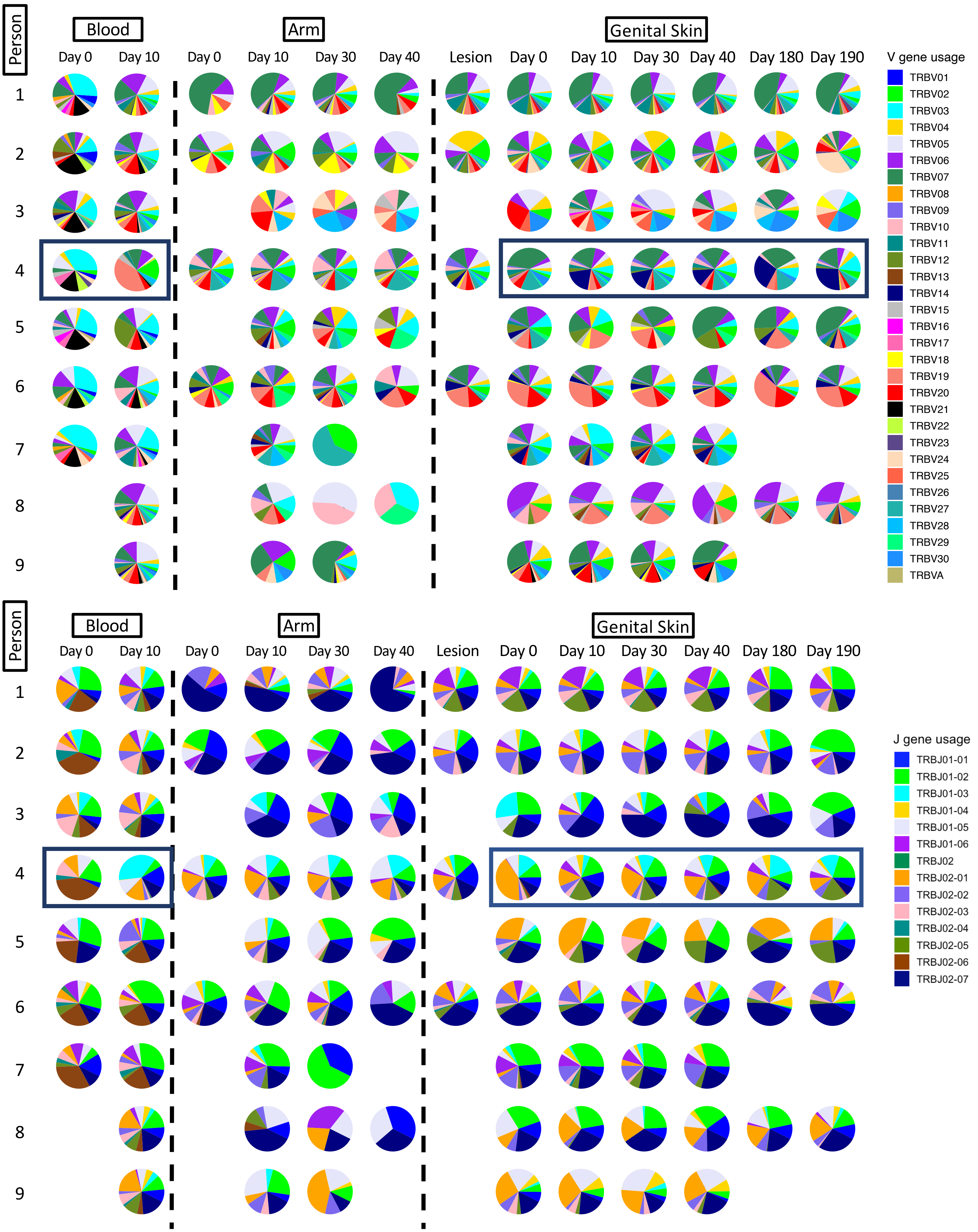

Supplementary Figure 4.

**a.**

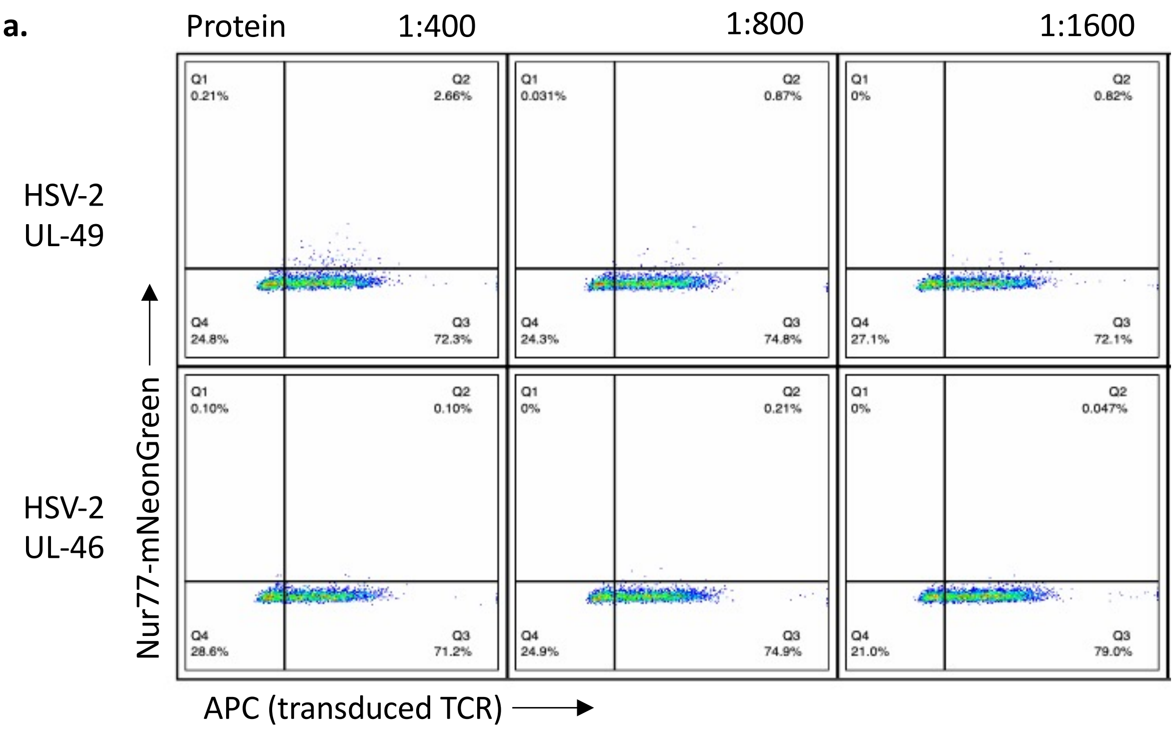


**b.**

81-93 ARPRRSASVAGSH

| | | Pool 9 | Pool 10 | Pool 11 | Pool 12 | Pool 13 | Pool 14 | Pool 15 | Pool 16 | Pool 17 | Pool 18 |
|---|---|---|---|---|---|---|---|---|---|---|---|
| | Pool 1 | 0.302 | 38.5 | 0.277 | 0.204 | 0.178 | 0.373 | 0.255 | 0.304 | 0.201 | 0.174 |
| 81-93 | Pool 2 | 35.8 | 53.7 | 32.9 | 35.5 | 36.1 | 37.1 | 36.2 | 37.1 | 37.7 | 38.2 |
| | Pool 3 | 8.21 | 41.7 | 5.25 | 5.56 | 5.95 | 5.97 | 7.75 | 6.62 | 6.89 | 7.82 |
| | Pool 4 | 0.174 | 27.1 | 0.148 | 0.226 | 0.127 | 0.2 | 0.241 | 0.255 | 0.211 | 0.259 |
| | Pool 5 | 0.358 | 32.3 | 0.25 | 0.257 | 0.126 | 0.101 | 0.16 | 0.233 | 0.0265 | 0.109 |
| | Pool 6 | 0.232 | 40.8 | 0.154 | 0.129 | 0.104 | 0.183 | 0.0802 | 0.0524 | | 0.27 |
| | Pool 7 | 0.0518 | 39.7 | 0.154 | 0.257 | 0.176 | 0.283 | 0.321 | 0.208 | | |
| | Pool 8 | 0.129 | 32.3 | 0.103 | 0.256 | 0.0796 | 0.157 | 0.214 | | | |

**c.**

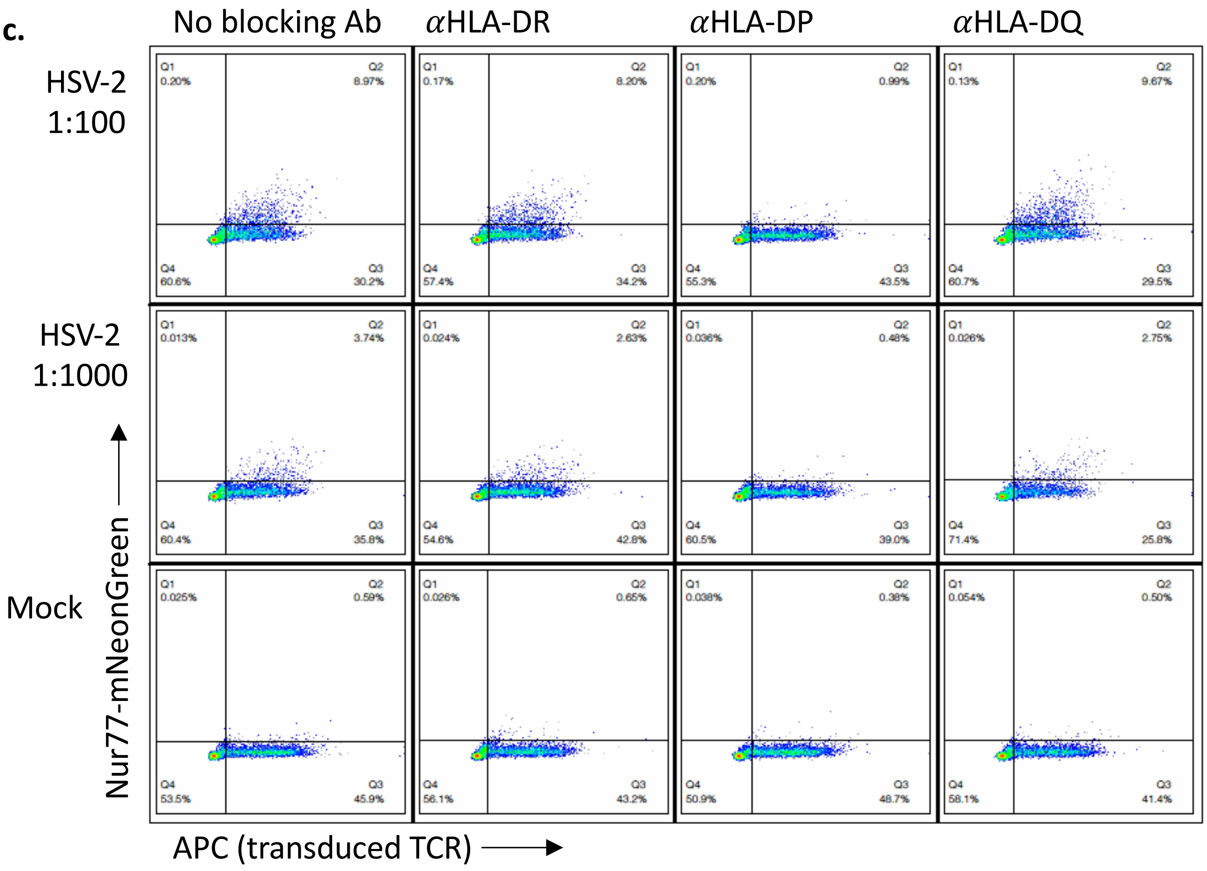

**Supplementary Figure 5.**

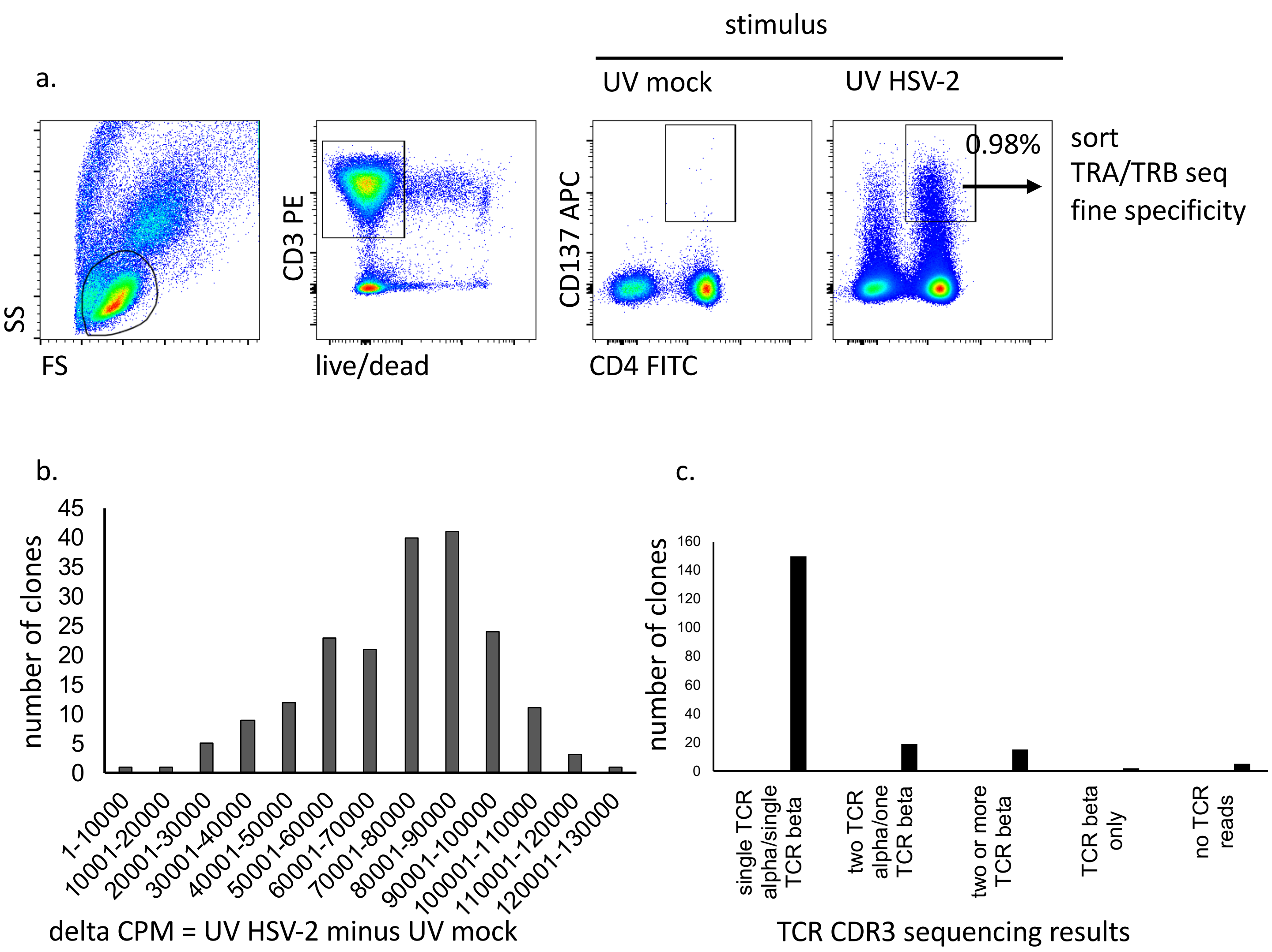
